# High resolution linear epitope mapping of the receptor binding domain of SARS-CoV-2 spike protein in COVID-19 mRNA vaccine recipients

**DOI:** 10.1101/2021.07.03.21259953

**Authors:** Yuko Nitahara, Yu Nakagama, Natsuko Kaku, Katherine Candray, Yu Michimuko, Evariste Tshibangu-Kabamba, Akira Kaneko, Hiromasa Yamamoto, Yasumitsu Mizobata, Hiroshi Kakeya, Mayo Yasugi, Yasutoshi Kido

## Abstract

The prompt rollout of the coronavirus disease (COVID-19) messenger RNA (mRNA) vaccine is facilitating population immunity, which shall become more dominant than natural infection-induced immunity. At the beginning of the vaccine era, understanding the epitope profiles of vaccine-elicited antibodies will be the first step in assessing functionality of vaccine-induced immunity. In this study, the high-resolution linear epitope profiles of Pfizer-BioNTech COVID-19 mRNA vaccine recipients and COVID-19 patients were delineated by using microarrays mapped with overlapping peptides of the receptor binding domain (RBD) of severe acute respiratory syndrome coronavirus 2 (SARS-CoV-2) spike protein. The vaccine-induced antibodies targeting RBD had broader distribution across the RBD than that induced by the natural infection. Thus, relatively lower neutralizability was observed when a half-maximal neutralization titer measured *in vitro* by live virus neutralization assays was normalized to a total anti-RBD IgG titer. However, mutation panel assays targeting the SARS-CoV-2 variants of concern have shown that the vaccine-induced epitope variety, rich in breadth, may grant resistance against future viral evolutionary escapes, serving as an advantage of vaccine-induced immunity.

**Importance:** Establishing vaccine-based population immunity has been the key factor in attaining herd protection. Thanks to expedited worldwide research efforts, the potency of messenger RNA vaccines against the coronavirus disease 2019 (COVID-19) is now incontestable. The next debate is regarding the coverage of SARS-CoV-2 variants. At the beginning of this vaccine era, it is of importance to describe the similarities and differences between the immune responses of COVID-19 vaccine recipients and naturally infected individuals. In this study, we demonstrated that the antibody profiles of vaccine recipients are richer in variety, targeting a key protein of the invading virus, than those of naturally infected individuals. Yet vaccine-elicited antibodies included more non-neutralizing antibodies than infection-elicited, their breadth in antibody variations suggested possible resilience against future SARS-CoV-2 variants. The antibody profile achieved by vaccinations in naive individuals pose important insight into the first step towards vaccine-based population immunity.

## Introduction

Messenger RNA (mRNA) vaccines have prevailed globally to mitigate the coronavirus disease 2019 (COVID-19) pandemic. Given the prompt progress in the development of vaccines and their fast rollout at global scale, population immunity against the severe acute respiratory syndrome coronavirus 2 (SARS-CoV-2) will largely depend on vaccine-induced rather than the infection-induced immunity. In this start of the COVID-19 vaccine era, the *de novo* repertoire of vaccine-elicited antibodies in SARS-CoV-2 infection-naive individuals will be the first step to build an optimal host defense system towards vaccine-based population immunity.

Currently, the efficacy of vaccine-induced immunity against SARS-CoV-2 in an individual is evaluated by potential surrogate markers such as half-maximal neutralization titers (NT50) using live/pseudo viruses and total antibodies titers against the receptor binding domain (RBD) of the spike protein of the virus (1–4). Understanding the epitope profile of both vaccine recipients and naturally infected individuals can readily help elucidating further molecular basis of these markers as surrogate. Moreover, the coevolution of vaccine-induced host immunity and the virus escape will be one of the most important elements to consider in the way of achieving herd immunity against COVID-19.

The RBD of the spike protein of SARS-CoV-2 is widely considered the key protein target for designing vaccines and developing neutralizing antibodies as therapeutic agents (5,6). Epitope profiles of sera from individuals naturally infected with COVID-19 have enabled to identify several immunodominant regions in the spike protein (7–9). While most immunodominant epitopes locate outside the RBD, the minor proportion targeting specifically the neutralizing RBD epitopes explain the majority of viral neutralizability and protection against re-exposures (10,11). In fact, neutralizing monoclonal antibodies (NAbs) developed as potential therapeutics also mainly target the epitopes located in the RBD (6,12–16). While a growing number of individuals acquire vaccine immunity, the detailed epitope profile of the humoral immune response to the mRNA vaccine is not fully understood (1,17,18).

In this study, high resolution linear epitope profiling targeting the RBD was performed using sera of both mRNA vaccine recipients and COVID-19 patients. By comparing the epitope profiles, we sought to describe the similarities and differences between the humoral immune responses induced by BNT162b2 mRNA (Pfizer/BioNTech) vaccination and natural infection. Information provided by this study will be crucial in this post-vaccine era of the COVID-19 pandemic.

## Materials and Methods

### Serum collection sufficient

Two groups were analyzed in this study: (i) vaccine recipients, all received two doses of BNT162b2 mRNA vaccine (Pfizer/BioNTech) with a three-week interval (N=21, age 20s–80s years old). Blood was obtained 17–28 days after the second dose. (ii) COVID-19 patients confirmed by nucleic amplification testing (N=20, age 20s–80s years old). The blood collection of the patients was performed between 10 and 63 days (median 39 days) after the onset. Detailed information of the subjects and severity of the disease of the patients can be found in Supplemental Table 1 (19).

Blood samples were obtained by venipuncture in serum separator tubes and the serum fraction was stored at – 80°C. All subjects provided written consent before participating in this study. This study was approved by the institutional review board.

### Anti-RBD IgG quantification by chemiluminescent immunoassay

Anti-RBD IgG titers of both groups were quantitated by measuring the chemiluminescence generated in the reaction mix containing serum IgG-bound, RBD-coated microparticles and acridinium-labeled anti-human IgG (Abbott SARS-CoV-2 IgG II Quant assay, USA)(20). Antibodies targeting the viral nucleocapsid protein (Anti-N IgG) were also measured for the sera of vaccine recipients to screen unrecognized exposure to SARS-CoV-2 (Abbott SARS-CoV-2 IgG assay, USA) (21).

### Neutralization assay using live SARS-CoV-2

The neutralization assay was carried out as described previously (22), but with modifications. Heat-inactivated (at 56°C for 45 minutes) vaccine-recipients and patients’ sera and a SARS-CoV-2 negative control serum were serially four-fold diluted with Dulbecco’s Modified Eagle Medium with 2% fecal bovine serum (2% FBS DMEM) and incubated with a pre-titrated 150 focus-forming units of SARS-CoV-2 JPN/TY/WK521 strain live virus particles (National Institute of Infectious Diseases, Japan) at 37°C for 1 hour. The monolayer of VeroE6 cells (National Institutes of Biomedical Innovation, Health and Nutrition, Japan) were then absorbed with the mixtures at 37°C. After a 1-hour incubation, the mixtures were replaced with fresh 2% FBS DMEM. After an 8-hour culture at 37°C, infection rates of the cells were determined by immunofluorescent staining, as follows. After fixation (4% paraformaldehyde, 15 minutes), cells were permeabilized (0.1% TritonX100, 15 minutes) and incubated with rabbit anti-spike monoclonal antibodies (Sino biological, China) (1:1000, 1 hour at 37°C). Cells were then washed and incubated with Alexa488-conjugated goat anti-rabbit IgG (Thermofisher scientific, USA) (1:500, 45 minutes at 37°C). Antigen positive cells were counted under a fluorescent microscope and the percentage of neutralization was estimated as the viral infectivity under serum-treated conditions compared with that without serum.

### Epitope mapping of the RBD

For precise linear epitope screening, overlapping 15-mer peptides (shift by 3 amino acids) were sequentially synthesized according to the sequence of the RBD on cellulose membrane by MultiPep synthesizer (Intavis Bioanalytical Instruments, Germany) using SPOT technology (23,24). The sequence of the RBD was obtained by GenBank (accession: MN908947.3, S319–S541) Additional 15-mer peptides containing single mutations of variants of concerns found within the RBD were designed. Single mutations included K417N, K417T, E484K and N501Y (25). Detailed peptide sequences used in this study can be found in Supplement Table 2.

Synthesized arrays were probed with sera at a 1:400 dilution followed by incubation with horseradish peroxidase conjugated goat anti-human IgA, IgG, and IgM polyclonal antibody at a 1:30,000 dilution. The bound of the secondary antibody on each peptide was detected and quantified by enhanced chemiluminescence. The peptide synthesis, probing and quantification were outsourced to Kinexus Bioinformatics Corporation. The epitopes were detected by subjective visual inspection. Our cutoff for signal detection was set at visually detectable peaks in a graph depicting a mostly minimum of 0.5 z-score of the mean peptide signals and/or regions previously reported as neutralizing antibodies in the RBD (27,28).

### Statistical analysis

Chemiluminescence signal intensities of the peptide arrays were standardized in two ways: relative values to the maximum signal level of each array as 100, and z-scores considering peptide signals of individual subjects as population. These calculations were done by Microsoft Excel for Microsoft 365 MSO (16.0.14026.20202).

Nonlinear regression curve fitting was performed to calculate half-maximal neutralization titers (NT50) of the neutralization assay. Statistical significance was calculated using un-paired two-tailed t-test. GraphPad Prism 9.1.0.221 was used for these statistical analyses.

The sequence and conformational information of the RBD was obtained under the accession number 6M0J (5) and 7A94 (26) at Protein Data Bank (PDB). The images to depict the recognized epitopes are shown using The PyMOL (Molecular Graphics System, Version 1.2r3pre, Schrödinger, LLC).

## Results

### Total IgG titers targeting the RBD and neutralization assay using live SARS-CoV-2

All vaccine recipients (N=21) and COVID-19 patients (N=20) revealed seropositivity to anti-RBD IgG according to the manufacturer’s threshold (>50 AU/mL) and the two groups did not show significant difference in their levels of anti-RBD IgG titers (Figure 1a). However, the neutralization assay using live SARS-CoV-2 showed remarkably lower NT50 in vaccine recipients compared to COVID-19 patients (p=0.0035) (Figure 1b). The ratio between the anti-RBD IgG antibody titer and the NT50 value was calculated in individual as shown in Figure 1c. It appeared that the anti-RBD IgG/NT50 ratios were significantly higher in vaccine recipients compared to COVID19 patients (p<0.001) (Figure 3c). The result indicated that the sera of vaccine recipients were more abundant in non-neutralizing, mere binding IgG antibodies, suggesting a discrepancy in the epitope profiles between vaccine recipients and COVID-19 patients. None of the vaccine recipients were seropositive to anti-N IgG, ensuring that they were naive to SARS-CoV-2 infection (Supplement Table 1).

**Figure 1.**
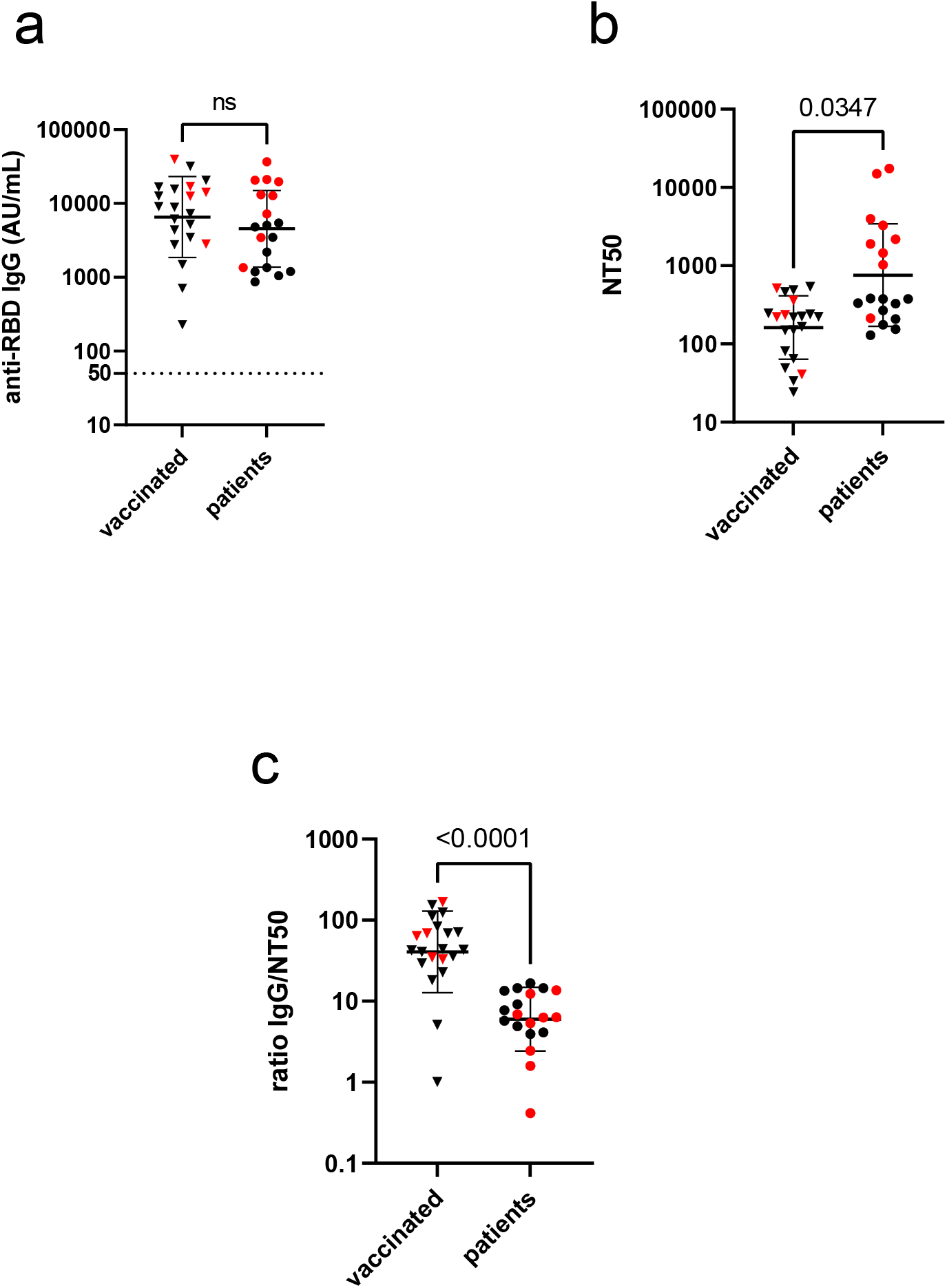
Total antibody titers targeting the RBD and neutralization of live SARS-CoV-2. (a) Anti-RBD IgG titers of vaccine (BNT162b2) recipients (N=21) and COVID-19 patients (N=19) were depicted. No significant difference in level of total anti-RBD IgG titers was observed. (b) The half-maximal neutralization titers (NT50) were remarkably lower in the vaccinated group than in patients. (c) Anti-RBD IgG/NT50 ratio was plotted in both groups. Black horizontal bars indicate geometric mean with geometric standard deviation. For detailed information on subjects, see Supplement Table1.

### Comparison of linear epitope profiles targeting the RBD of vaccine-elicited and infection-elicited sera

To delineate the discrepancy in the epitope profiles between vaccine recipients and COVID-19 patients with high resolution, we next mapped and compared the immunodominant epitopes of both sera by using an overlapping 15-mer linear-peptide array (Figure 2a). Sera of vaccine recipients and COVID-19 patients were incubated with the microarray. Sera of five subjects from the vaccine recipients and ten from the patients were selected based on their anti-RBD antibody titers and NT50 (denoted as red dots in Figure 1). The sera were incubated with the designed microarrays arranged with 15-mer overlapping peptides of the RBD on the surface (Figure 2a). The designated array did not show any considerable unspecific binding of the secondary antibody.

**Figure 2.**
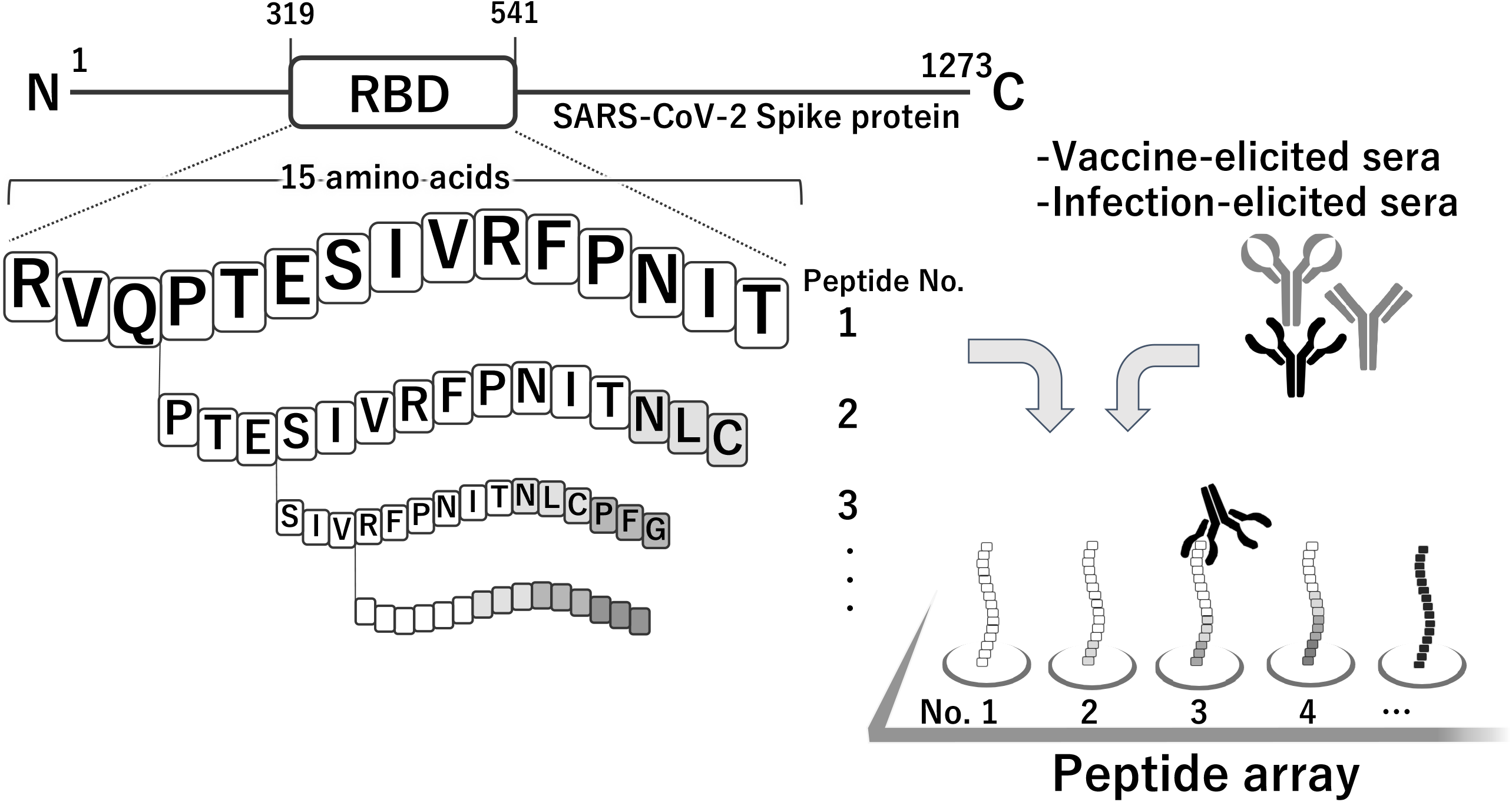
High resolution linear epitope mapping of the receptor binding domain (RBD) of the SARS-CoV-2 spike protein. (a) Overlapping 15-mer peptides (shift by 3 amino acids) of the RBD was sequentially synthesized on cellulose membrane. Sera of vaccine recipients and COVID-19 patients were incubated with the microarray, followed by the procedure mentioned in the methodology section to detect the reactive peptides. (b) Heat map identifying peptides recognized by IgG, IgA, and IgM in sera of vaccine recipients (Sample A–E) and COVID-19 patients (sample F–O). Signal of each peptide was calculated to relative value to the maximum signal of each subject as 100. Legend shows the darker the blue gets; the more signal was observed at the designated peptide.

We generated a heatmap according to the relative signals of the overlapping peptides (Figure 2b). Also, z-scores of each peptide were compared individually (Supplement Figure 1) and per group (Figure 3). Comparing the epitope profiles of the two groups, two types of epitopes were identified: (1) epitopes recognized by both groups and (2) epitopes recognized only by vaccine-elicited sera. Overall, seven linear epitopes were recognized within the RBD, four within (1): T415–F42, peptide No.33; R457–S477, peptide No.47–49; V433–N450, peptide No.39–40; V395-A411, peptide No.26– 27 and three within (2): N334–A348, peptide No.6; S373–L390, peptide No.19,20; S514–F541, peptide No.66–71, respectively (Figure 2b, Figure 3).

**Figure 3.**
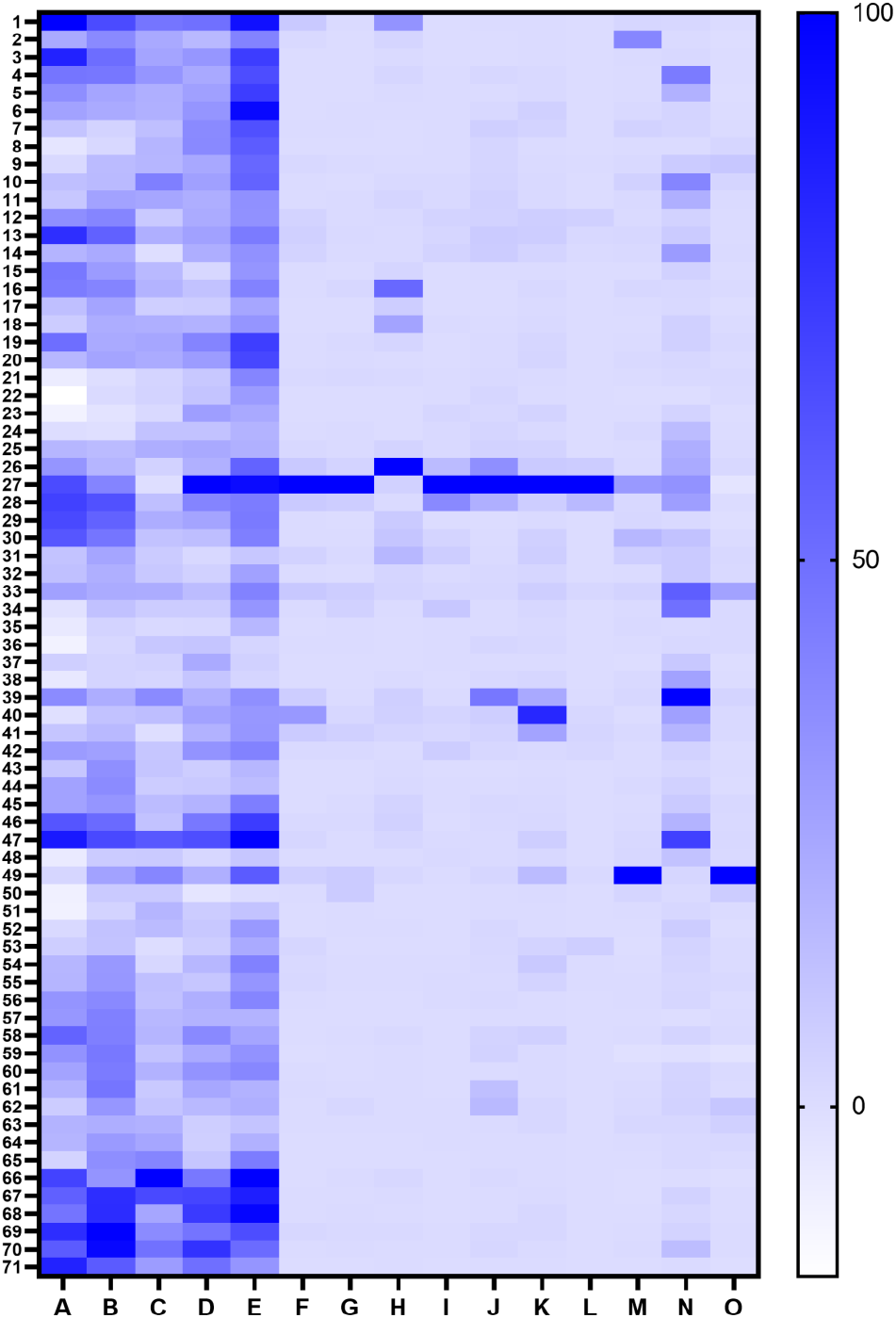
Comparison of epitope profiles between two groups: BNT162b2 vaccine recipients (N=5) and COVID-19 patients (N=10). (a) Thin red lines denote peptide signals of individuals. Bold red lines depict the mean values of the peptide signals of the COVID-19 patients’ sera (N=10). (b) Thin grey lines denote peptide signals of individuals. Bold black lines denote the mean values of the peptide signals of the vaccine recipients’ sera (N=5). (c) Red arrows denote epitopes recognized in the sera of both groups. Black arrows denote epitopes identified only in the vaccine recipients’ sera. Designated peptide numbers are shown above the arrows.

### (1) Epitopes recognized by both groups

A total of four linear epitopes were recognized in both groups (Figure 2b, Figure 3c). Three (peptide No.33, No.39–40 and No.47–49) of them shared the epitope regions of the RBD with neutralizing monoclonal antibodies previously reported as class1 and class 3 (27).

Linear epitopes were identified at peptide No.33 and peptides No.47–49 (Figure 3, Figure 4a,b), sharing the epitopes with reported class 1 neutralizing antibodies (27). Also, peptide No.39–40 (Figure 3, Figure 4c) shared epitope residues very similar to human monoclonal antibody REGN10987, categorized as class 3 neutralizing antibody which sterically hinders the interaction between angiotensin converting enzyme 2 (ACE2) and the RBD (27,29).

**Figure 4.**
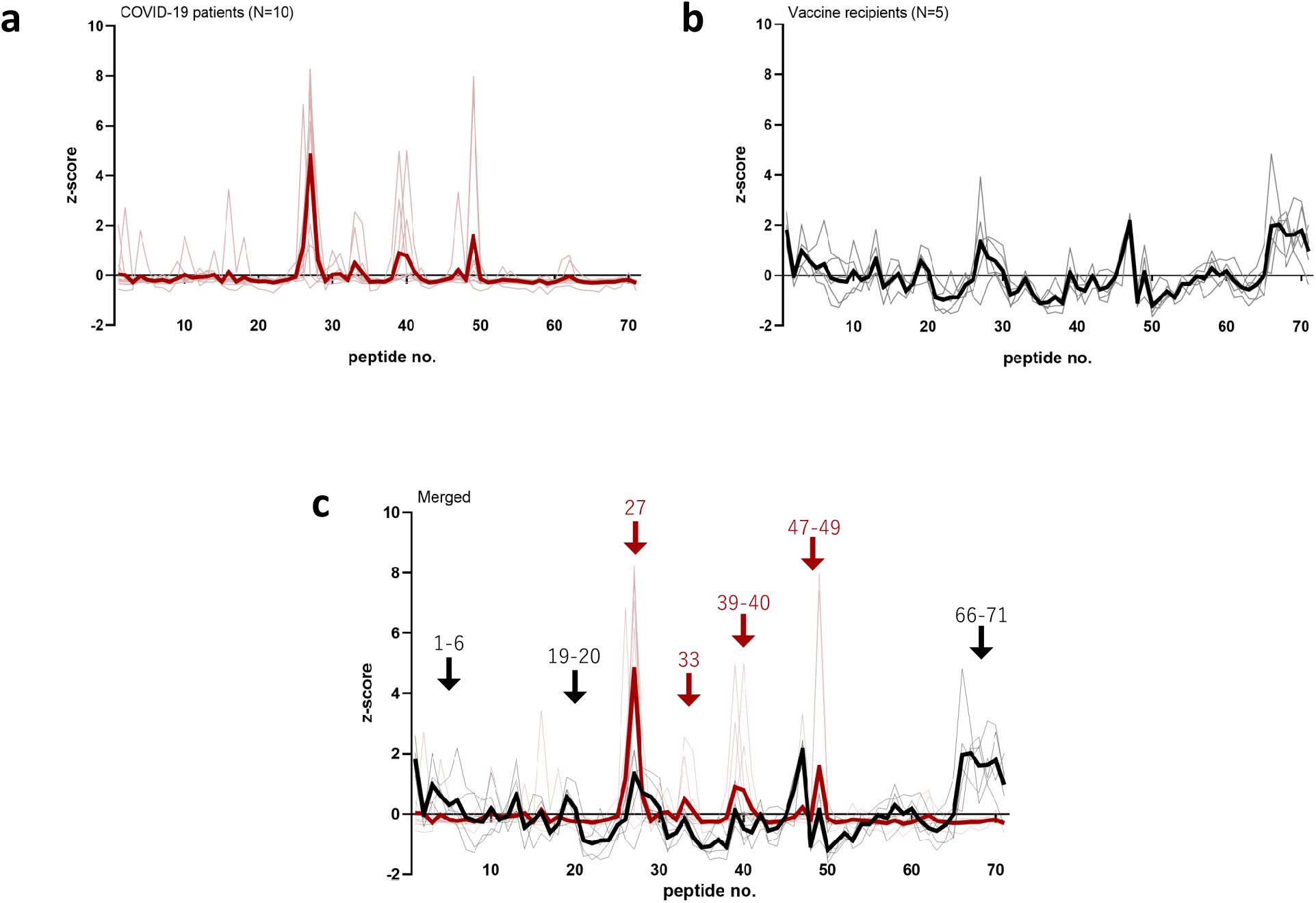
Linear epitopes in the receptor binding domain (RBD) identified in both groups of vaccine recipients and COVID-19 patients. Angiotensin converting enzyme2 (ACE2); green, the RBD; tint blue, (a) linear epitope T415-F429 (peptide No.33); cyan (b) linear epitope R457-S477 (peptide No.47-49); yellow (c) linear epitope A433-S450 (peptide No.39-40); blue.

The linear epitope, peptide No.26–27, was reactive at the highest level in most serum samples, as sera of 2/5 vaccine recipients and 7/10 patients had maximum reactivity to this peptide (Figure 3c, Supplement Figure 1). However, antibodies binding to this epitope seemed not to contribute to neutralizing the live virus, based on our observations detailed below. This peptide was found to be equally reactive, at a high extent, to a serum with negligible neutralizability, obtained from a COVID-19 patient who had undergone Rituximab treatment (Details found in Supplement Figure 2) (30). In the RBD structure, the No.27 peptide is located inside the core β sheets, which is not exposed to the surface of the RBD in either an “up” or “down” position. Judging from the structural composition, this linear epitope would not affect ACE2 binding (Supplement Figure 2).

### (2) Epitopes recognized only in vaccine recipients’ sera

Three linear epitopes of the RBD were uniquely found in vaccine recipients’ sera (Figure 2b, Figure 3b,c), two of them (peptide No.6 and No.19,20) were sharing the epitope regions of the RBD with neutralizing monoclonal antibodies known as class 3 and class 4 (27).

At N-terminus of the RBD, namely the peptide No.1–6, we identified an epitope region detected only in the post-vaccination sera (Figure 2b, Figure 3c). Especially, peptide No. 6 was an epitope of note, which shall be recognized by the class3 NAb S309 by P337–A344 helix residues (Figure 5a) (31). The epitope is distinct from the receptor-binding motif and has a good accessibility both in the up and down compositions of the RBD (PDB, 7A49, Figure 5b).

**Figure 5.**
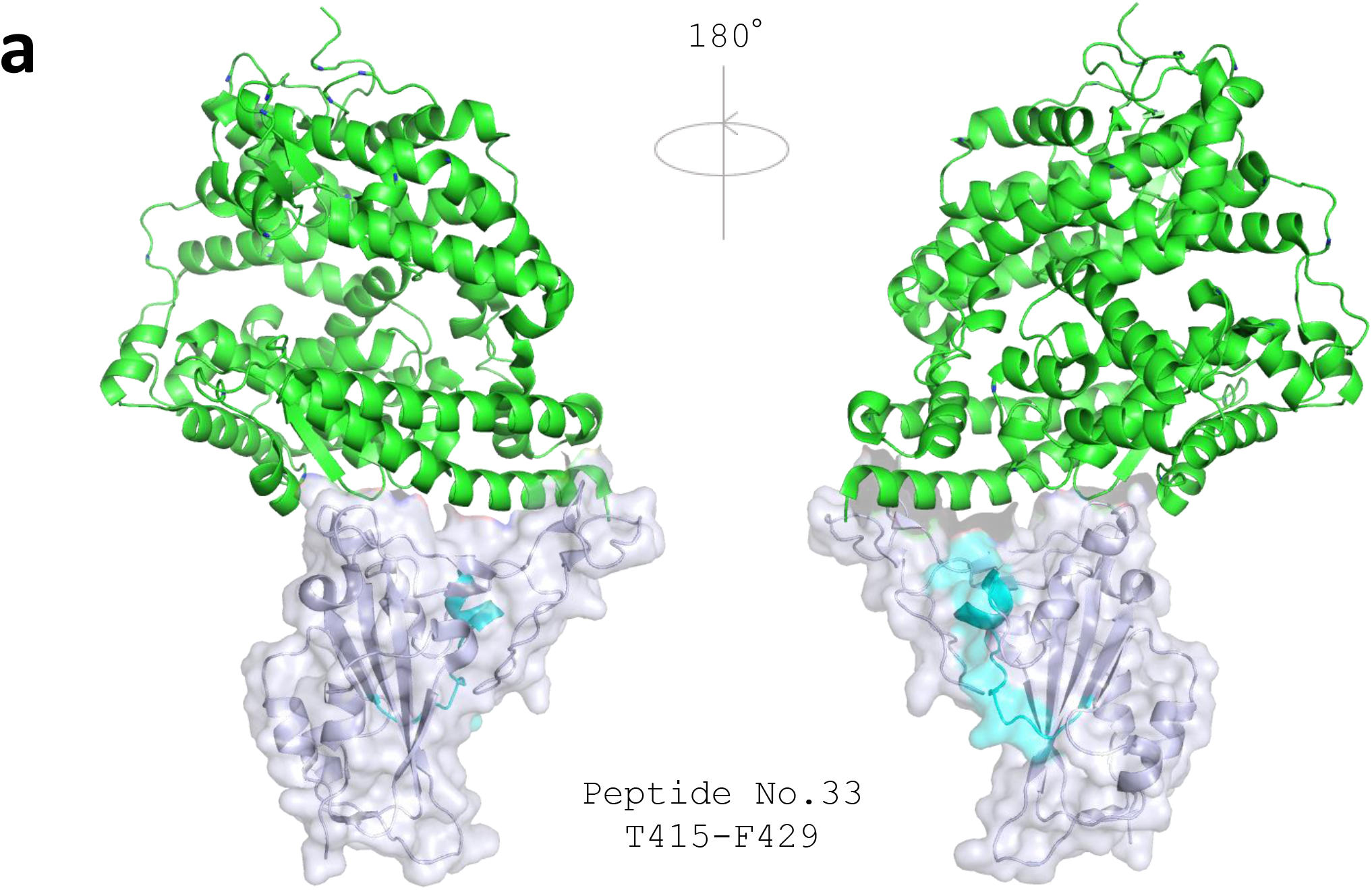
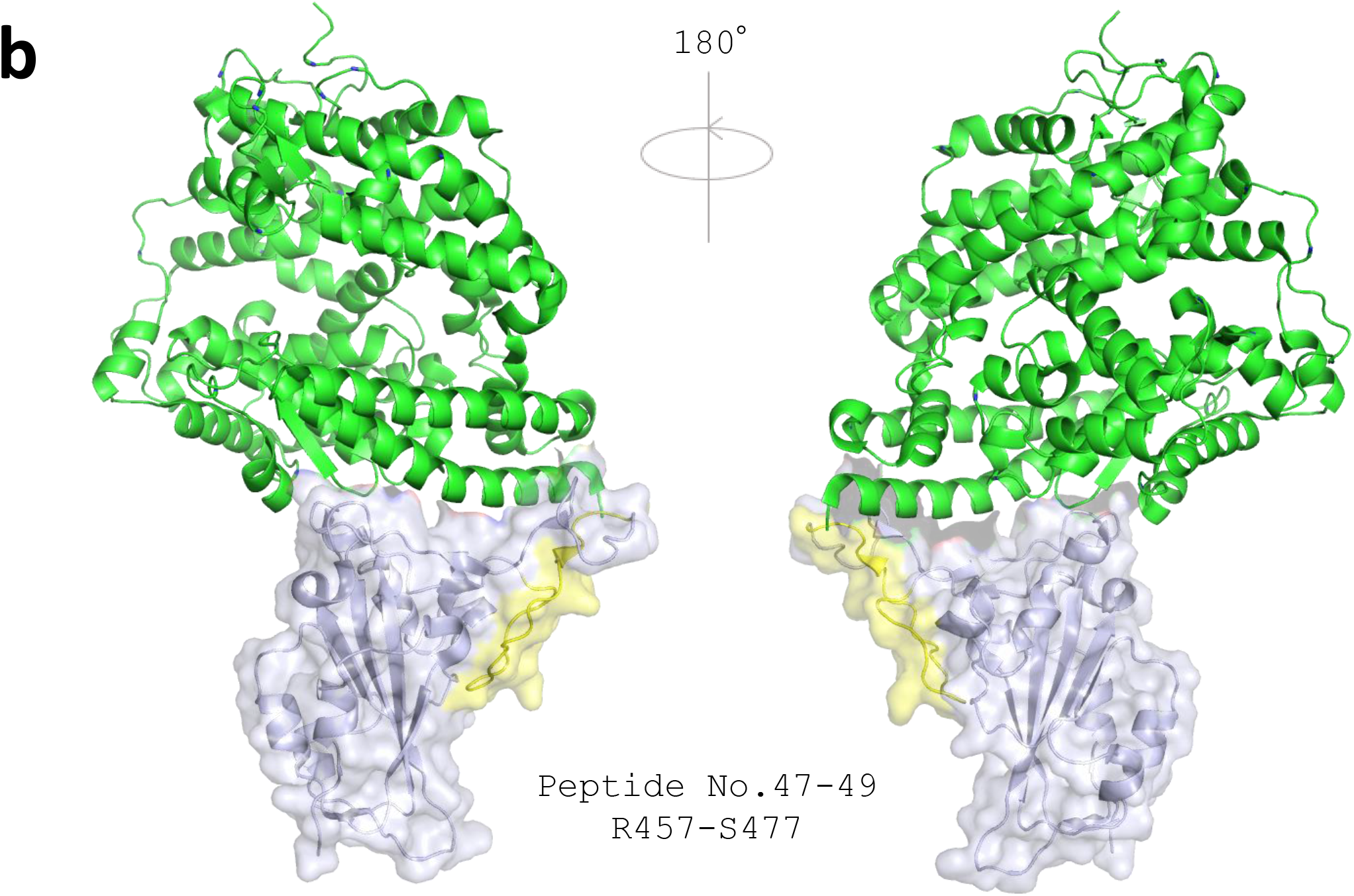
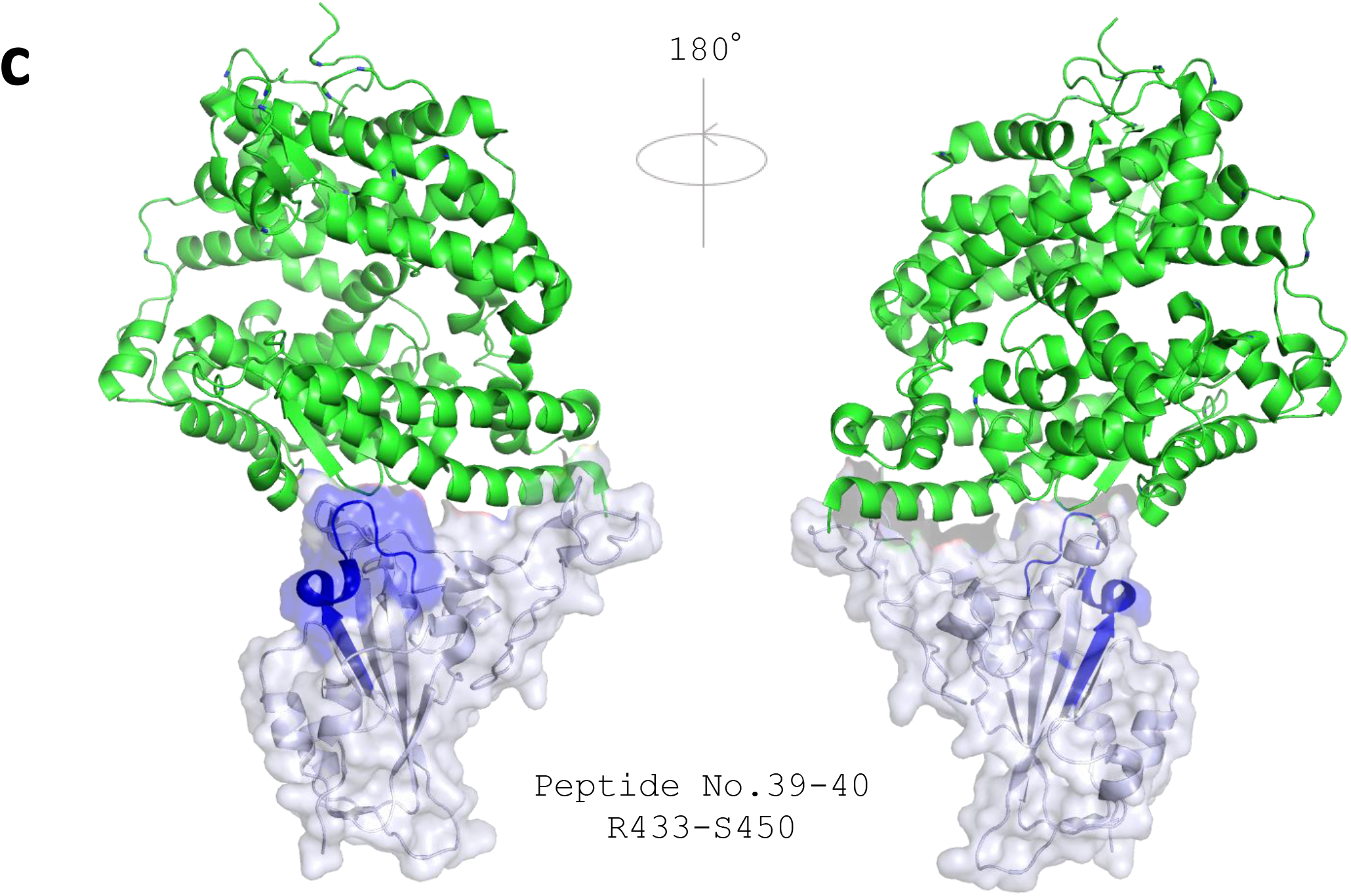
Linear epitopes in the receptor binding domain (RBD) identified only in vaccine recipients’ sera. Angiotensin converting enzyme2 (ACE2) is shown in green. (a) The RBD is shown in tint blue. Linear epitope R334-S348 (peptide No.6); magenta. (b) SARS-CoV-2 spike trimer in one open, two closed (one RBD up, two RBD up) composition. Spike subunit 2 and N-terminal domain are in the same color, light blue, yellow, and tint blue. Up RBD is in dark blue, Down RBDs are in yellow. Linear epitope R334-S348 has good accessibility in both up and down composition of the RBD. (c) The RBD; green. Linear epitope S373-L390 (peptide No.19-20); orange.

Another identified epitope, peptide No.19 and 20, shared epitope residues with a neutralizing monoclonal antibody CR3022, categorized as class 4, isolated from a SARS-CoV convalescent (32,33). This class 4 neutralizing antibody attaches to the RBD but distal to the ACE2 binding site and is highly conserved among different CoV species (34).

The third epitope, located at the peptide No.66–71, did not match with any known monoclonal antibodies. Yi et al. detected the same region of the peptides (V524–F541) reactive from COVID-19 convalescent serum in their linear epitope analysis (11). They also demonstrated that these peptides interacted with control sera as well (11). Thus, we considered our results on the corresponding peptides to be non-specific.

### Linear epitopes mapping with single mutations found in SARS-CoV-2 variants

Our analysis included single amino acid mutations of the RBD that are reported in the SARS-CoV-2 variants of concerns including B.1.1.7, B.1.351 and P.1 (PANGO lineage (35)). Additional 15-mer peptides with the substituting amino acid, K417N, K417T, E484K, and N501Y, were incubated with both vaccine recipients’ and patients’ sera. Interestingly, vaccine-induced sera showed consistent signals to the mutated peptides, whereas patients’ sera had almost no reaction (Figure 6).

**Figure 6.**
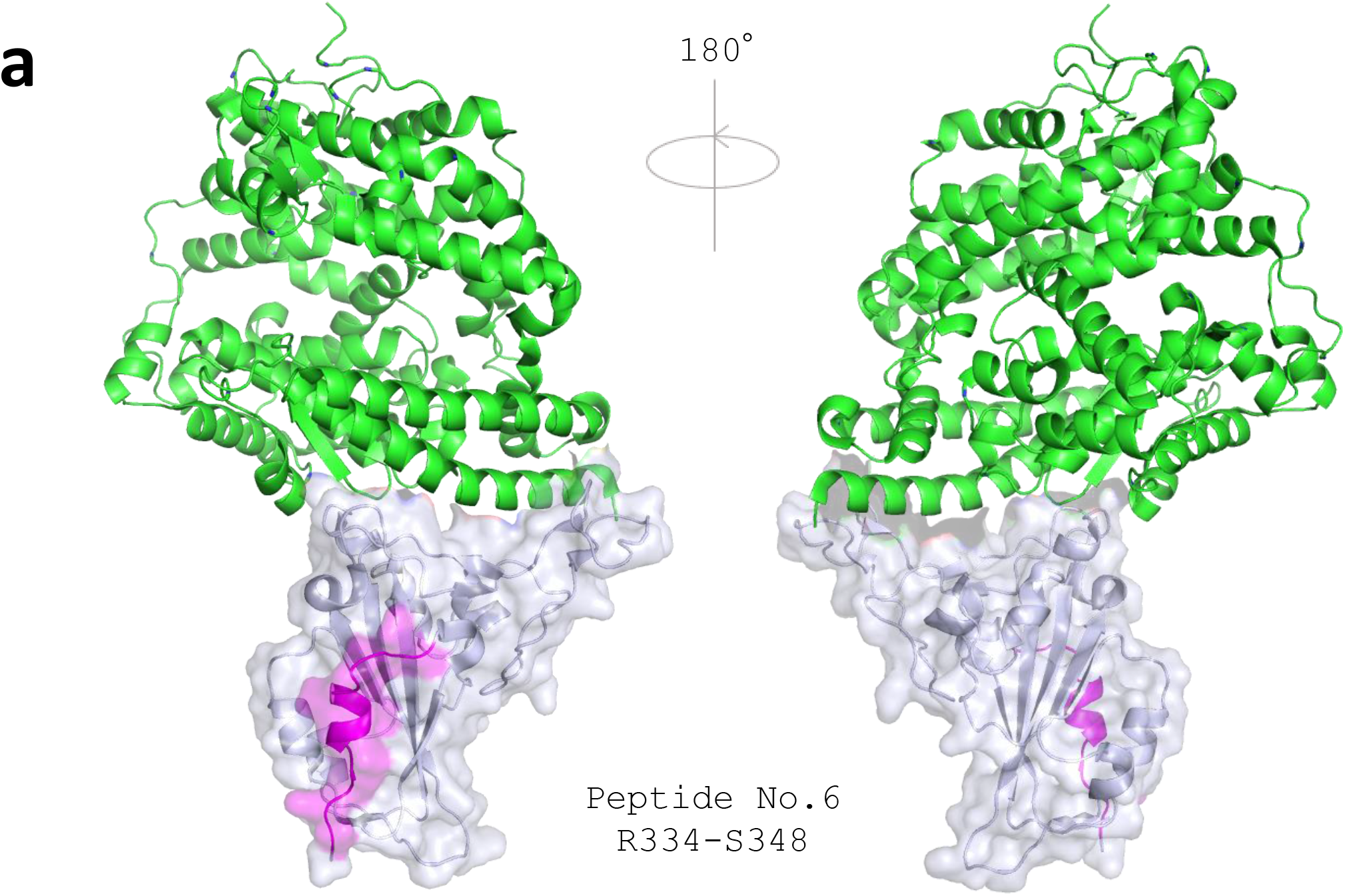
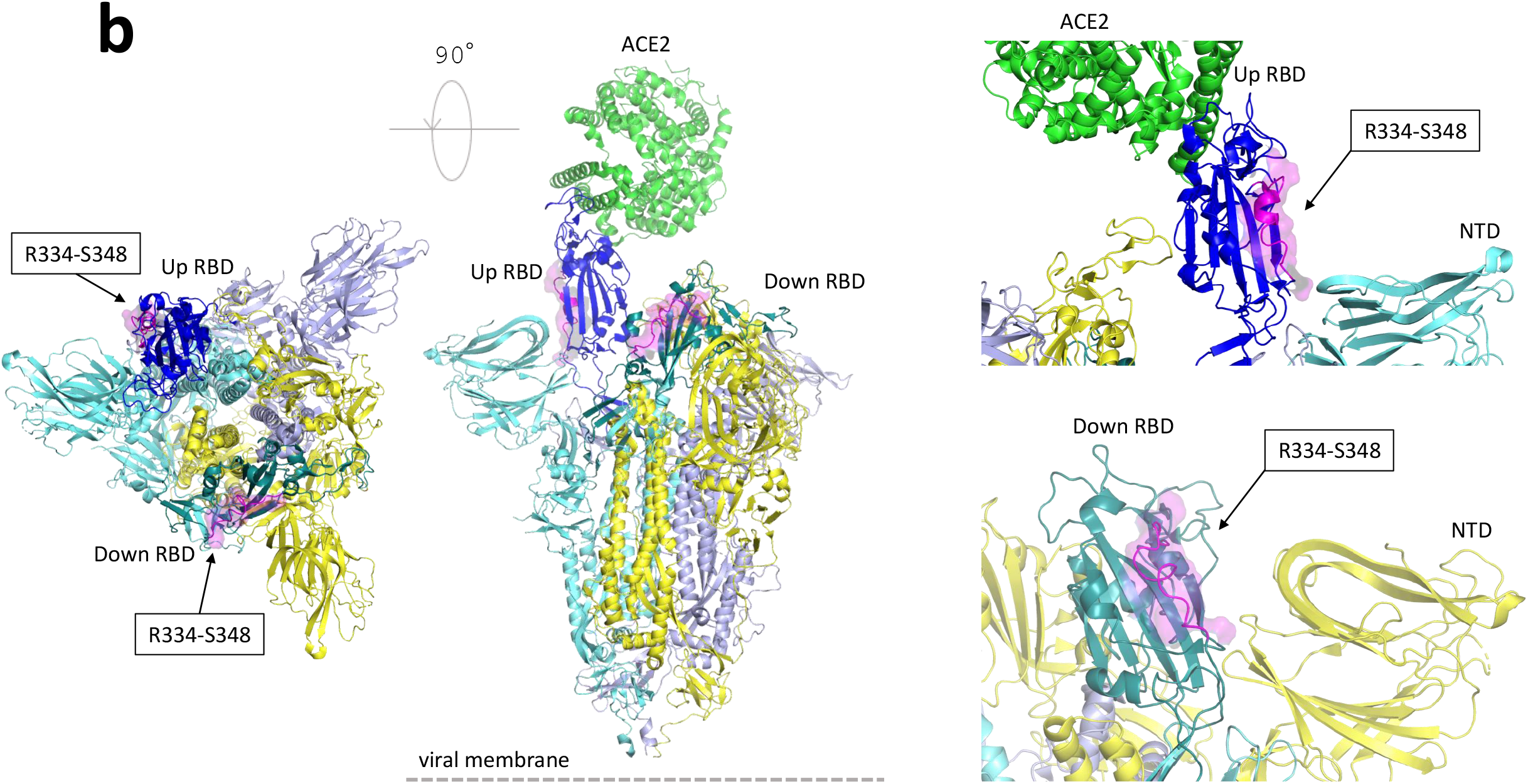
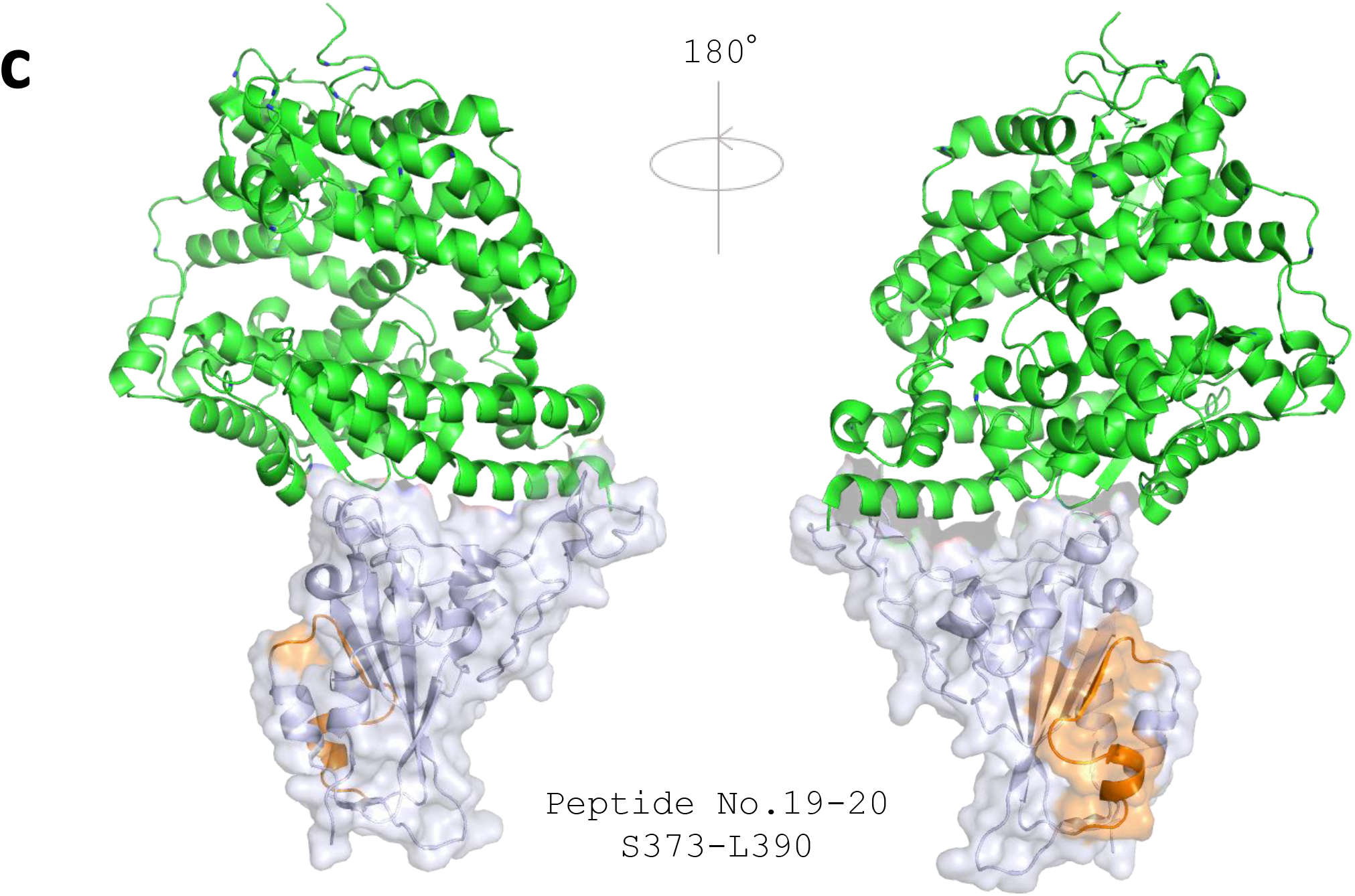
Mutation peptide panels showed more reactivity to vaccine recipients’ sera than to patients’ sera. Heat maps identifying peptides encoding the single mutations, recognized by IgG, IgA, and IgM in sera of vaccine recipients (Sample A–E) and COVID-19 patients (sample F–O). Signal of each peptide was calculated to relative value to the maximum signal of each subject as 100.

In our original peptide array (Figure 2b, Figure 3), the overlapping peptides encoding the mentioned mutations sites (K417, E484, and N501 of the RBD) corresponded to peptides No.29–33, No.52–56, and No.57–61, respectively. The peptides No.52–61, containing E484 and N501, did not show any significant reaction in both vaccine recipients and COVID-19 patients (Figure 2b, Figure 3). The peptides No.29–33 containing K417, considered as escape mutation, was reactive to antibodies in both patients and vaccine recipients’ sera as mentioned above (Figure 2b, Figure 3).

## Discussion

This study revealed the linear epitope profiles targeting RBD elicited by BNT162b2 mRNA vaccination and natural infection of SARS-CoV-2. Our principal finding was that the variation of linear epitopes was broader in vaccine-elicited antibodies compared with infection-elicited antibodies, whichmay contribute to potent neutralization and thus resistance of the vaccine-elicited antibodies against the SARS-CoV-2 variants of concern.

Now, four categories of NAbs classes are proposed to characterize the mode of recognition and epitope specificity (27). Class 1 NAbs block several proximal sites in the receptor binding motif (RBM) of the RBD and directly block ACE2 binding (27); class 2 NAbs recognize both up and down formations of the RBDs and epitope overlapping or close to ACE2-binding site (27); class 3 NAbs recognize both up and down RBD and bind outside ACE2-binding site (27,31); class 4 NAbs bind only to up RBDs and do not directly block ACE2 binding, but destabilizes the virus’ prefusion spike conformation (33,34). Many of the human-isolated NAbs target RBD, while some target N-terminal domain of subunit 1 spike protein (14,36). In our study, two classes of NAbs exclusively relevant in vaccine-elicited sera were found to be of specific note; peptide No.6 (Figure 5a,b) targeted by the class 3 NAb (31) and peptide No.19–20 (Figure 5c) targeted by the class 4 NAb (33,34). These epitopes locate outside the ACE2-binding RBM (Figure 5), while epitopes commonly detected in both vaccine and infection-elicited repertoires clustered adjacent to the ACE2-binding site (Figure 4).

The majority of NAbs targeting the RBM, which correspond to class 1 and 2, have been shown to exhibit decreased neutralization against the virus variants (15,18,37). For example, antibodies recognizing the linear epitope, here included in peptide No.33, would possibly fail in neutralizing variants with K417N mutation as previously described (15,18,37,38). To the contrary, the linear peptides No.6 and No.19–20 (Figure 5a,b,c), corresponding to epitopes found exclusively in vaccine-elicited sera (Figure 2b, Figure 3c), revealed corresponding epitopes targeted by human NAbs isolated from SAR-CoV convalescent (S309 and CR3022, respectively) (31,33,34). These cross-neutralizing antibodies, belonging to class 3 and 4, recognize linear epitopes highly conserved among different CoV species. The epitopes recognized by these class 3 and class 4 NAbs are major contributors broadening the repertoire of vaccine-induced immunity. Located remotely from the RBM, such neutralizing epitopes stay rather free from variants (15,18), and explain the resistance of vaccine-elicited sera towards viral mutational escapes (39,40). The vaccine recipients’ broader epitope profile spanning across the RBD may give immunological flexibility and resilience against this evolving virus. Our mutation peptide panels have also presented a rather optimistic view on discussing the efficacy of vaccine-induced immunity to efficiently recognize the SARS-CoV-2 variants (Figure 6). However, considering that the linear epitope profiles harboring the mutation loci were not dominant in either vaccine sera or patients’ sera (Figure 2b, Figure 3b,c) and abundancy of conformational epitopes found adjacent to the RBM, the extent to which these specific linear epitopes contribute in net neutralizability remains yet to be determined (10,11,14,17).

We observed discrepancy between the neutralizability of sera obtained from vaccine recipients and patients, which could be partially explained by the difference in the time course of epitope selection and immune maturation. When comparing convalescent sera and vaccine-elicited immunity, the distribution of neutralizing epitopes was less generalized and focalized at specific peptides (Figure 3, Individual epitope distribution can be found in Supplement Figure 1). Among the two modes of acquired immunity, our results indicate that infection-induced humoral immunity had established a more mature, finely selected antibody repertoire. Our snapshot observations are in line with the ideas that maturation of infection-provoked repertoires occurs as early as 10–20 days after onset, or even earlier in the case of COVID-19 beginning at 4–7 days after onset (41,42). Positive selection of relevant epitopes and maturation of antibody repertoire thus may lag behind regarding the vaccine-induced immunity. Nevertheless, in this study, sera were sampled during the peak period of immune reaction in the host for both groups. Longitudinal evaluation of the epitope profiles and serological markers are needed to assess the further host immune evolution and draw conclusions to the above speculations.

In conclusion, we evaluated the similarity and difference in humoral immunity elicited by both BNT162b2 mRNA vaccine and natural infection of SARS-CoV-2. High resolution linear epitope profiles revealed the characteristic distribution of polyclonal antibodies spanning across the RBD in vaccine recipients’ sera, which possibly accounted for the discrepancy observed in serological markers. Based on the multiplicity of neutralizing epitopes supporting the protectivity of vaccine-elicited antibodies, mRNA vaccine-elicited humoral immunity may harbor advantages in resisting against the rapidly evolving pathogen.

### Limitations

There are several limitations in our study. The severity of the COVID-19 patients evaluated in this study was high (seven out of ten were critical) with comorbidities, whereas the vaccine recipients were relatively healthy without major comorbidities. The age was distributed in both groups. This analysis was focused exclusively on the linear epitope profile targeting RBD. Experimental observations on compositional epitopes nor epitopes outside the RBD region was not made in this study. Nonetheless, our results reporting the mRNA vaccine’s broader RBD epitope variety are in concordance with preceding reports (39,43).

## Data Availability

The data that supports the findings of this study are available in the supplementary material of this article.

## Author Approval

All authors have read and approved the manuscript.

## Acknowledgements

This work was funded by Japan Agency for Medical Research and Development (AMED) under Grant number JP20wm0125003 (Yasutoshi Kido), JP20he1122001 (Yasutoshi Kido), JP20nk0101627 (Yasutoshi Kido), and JP20jk0110021 (Yu Nakagama). This work was also supported by JSPS KAKENHI Grant Number JP21441824 (Natsuko Kaku). We received support from Osaka City University’s “Special Reserves” fund for COVID-19. We also receive the COVID-19 Private Fund (to the Shinya Yamanaka laboratory, CiRA, Kyoto University). Yuko Nitahara receives BIKEN Taniguchi Scholarship.

We are grateful for the virus provided by National Institute of Infectious Diseases, Tokyo, Japan. We appreciate James A. Rankin for his contribution in checking the manuscript.

## Author contribution

Nitahara Y, Nakagama Y, Kaku N and Kido Y designed the study.

Nitahara Y, Nakagama Y, Kaku N, Yamamoto H, Mizobata Y, Kakeya H, and Kido Y selected patients and acquired clinical data.

Nitahara Y, Nakagama Y, Kaku N, Candray K, Michimuko Y, Tshibangu-Kabamba E and Yasugi M performed immunological assays.

Nitahara Y, Nakagama Y, Kaku N and Kido Y performed epitope mapping analysis. Nakagama Y and Yasugi M performed neutralization assays.

Nitahara Y, Nakagama Y, Kaku N and Kido Y wrote the manuscript and contributed to analysis and interpretation of the data.

Yamamoto H, Mizobata Y, Kakeya H, Kaneko A and Yasugi M contributed to critical discussion of the manuscript.

**Supplement Figure 1.**
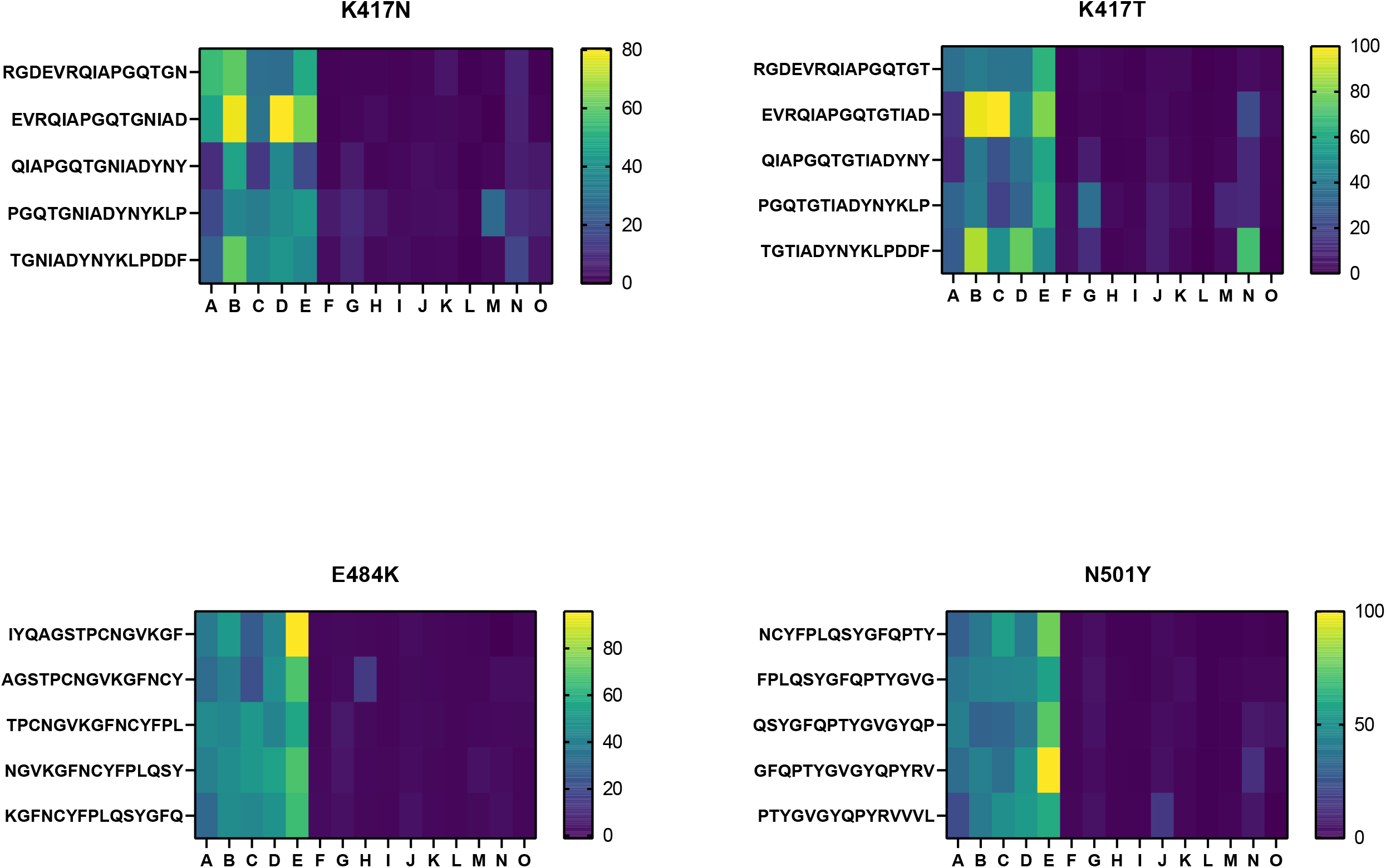
Epitope profiles of individual subjects are shown in graphs depicting z-scores of each peptide signal, calculated individually, on y-axis, overlapping peptide sequence on x-axis. Vaccine-induced sera (V01–V05) had more variety in recognizing epitopes than infection-induced sera (P01–P10).

**Supplement Figure 2.**
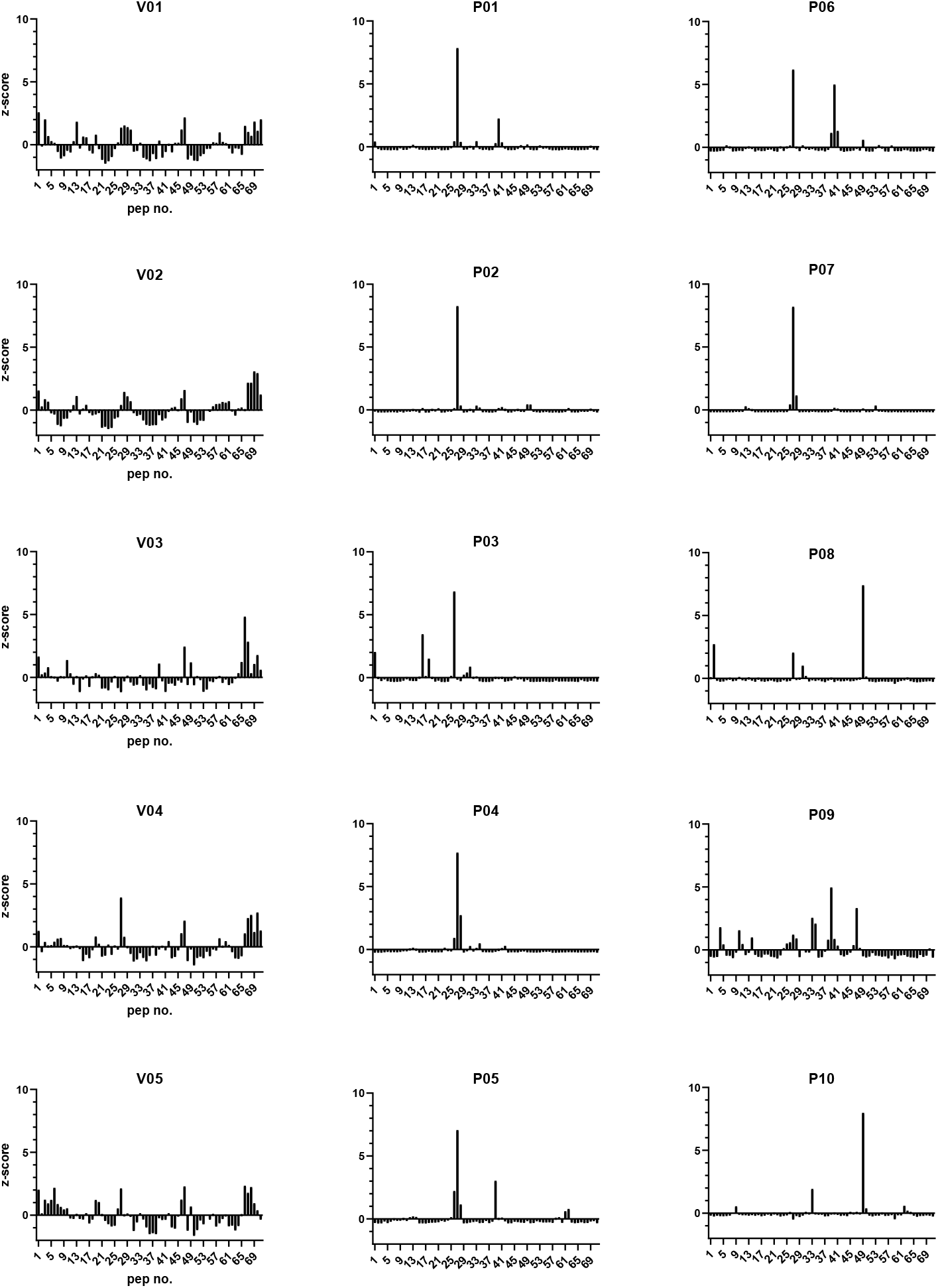
Epitope profile of a COVID-19 patient who had received Rituximab treatment. The antibodies targeting the peptide No. 27, corresponding to A397–A411 of RBD, were dominant in the epitope profile of this patient (depicted in the upper right graph), which showed limited neutralization compared to a COVID-19 positive serum sample from another COVID-19 patient. Graphic on the left shows the ACE2-RBD complex (ACE2 in green, RBD in tint blue). The position of peptide No.27 is depicted in dark gray.

**Supplement Table 1**. Detailed information of the subjects included in this study.

**Supplement Table 2**. Sequence of the peptides on microarrays used in this study.

**Supplement Table 3**. Raw signal of the microarrays.

---

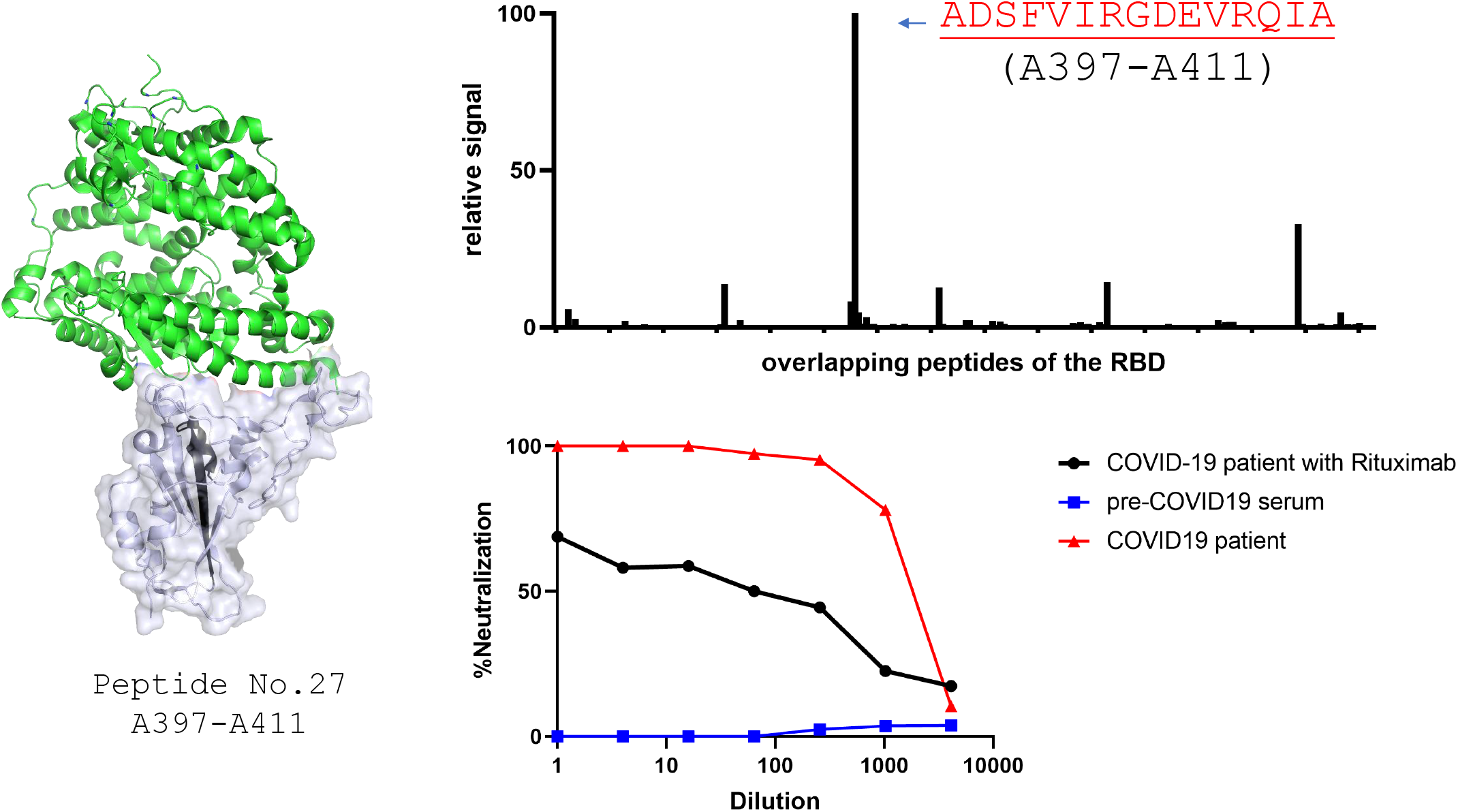

