## Supplement Table 1. for "High resolution linear epitope mapping of the receptor binding domain of SARS-CoV-2 spike protein in COVID-19 mRNA vaccine recipients"

Supplemental Table 1. Detailed information of subjects from vaccine recipients(N=21) and COVID-19 patients (N=20).

| Vaccine recipients | age range | sex | serum collection date after second dose (d) | NT50 | anti-RBD IgG | ratio anti-RBD IgG/NT50 | anti-N IgG |
|---|---|---|---|---|---|---|---|
| V01 | 31-40 | M | 21 | 151.1 | 12630.5 | 83.6 | 0.11 |
| V02*1 | 61-70 | M | 21 | 41.0 | 2829.3 | 69.0 | 0.23 |
| V03 | 31-40 | F | 21 | 463.6 | 31933.3 | 68.9 | 0.12 |
| V04 | 31-40 | M | 20 | 246.4 | 8870.4 | 36.0 | 0.06 |
| V05*1 | 51-60 | F | 19 | 235.6 | 39681.9 | 168.4 | 0.05 |
| V06*1 | 31-40 | M | 20 | 222.1 | 14224.0 | 64.0 | 0.06 |
| V07*1 | 31-40 | F | 20 | 360.3 | 12494.9 | 34.7 | 0.03 |
| V08*1 | 31-40 | F | 17 | 515.6 | 16998.9 | 33.0 | 0.40 |
| V09 | 31-40 | F | 21 | 218.8 | 15608.2 | 71.3 | 0.01 |
| V10 | 41-50 | M | 21-28 | 33.8 | 5234.3 | 154.8 | 0.03 |
| V11 | 41-50 | F | 21-28 | 165.7 | 7298.5 | 44.0 | 0.01 |
| V12 | 31-40 | F | 21-28 | 487.9 | 20656.3 | 42.3 | 0.04 |
| V13 | 51-60 | F | 21-28 | 49.3 | 6136.1 | 124.6 | 0.03 |
| V14 | 51-60 | F | 21-28 | 64.9 | 1485.3 | 22.9 | 0.02 |
| V15 | 51-60 | F | 21-28 | 222.9 | 9074.9 | 40.7 | 0.02 |
| V16 | 41-50 | F | 21-28 | 80.8 | 3484.7 | 43.2 | 0.02 |
| V17 | 31-40 | F | 21-28 | 24.2 | 710.1 | 29.3 | 0.04 |
| V18 | 41-50 | F | 21-28 | 539.9 | 2760.8 | 5.1 | 0.01 |
| V19 | 51-60 | F | 21-28 | 239.0 | 4371.4 | 18.3 | 0.01 |
| V20 | 31-40 | F | 21-28 | 149.7 | 16783.7 | 112.1 | 0.03 |
| V21 | 81-90 | F | 21-28 | 225.4 | 227.6 | 1.0 | 0.03 |

| COVID-19 patients | age range | sex | days after onset (d) | NT50 | anti-RBD IgG | ratio anti-RBD IgG/NT50 | severity of the disease*2 |
|---|---|---|---|---|---|---|---|
| P01*1 | 51-60 | M | 19 | 17414.0 | 7231.4 | 0.4 | critical |
| P02*1 | 31-40 | M | 12 | 1902.0 | 13110.0 | 6.9 | critical |
| P03*1 | 71-80 | M | 20 | 1449.0 | 19727.0 | 13.6 | critical |
| P04*1 | 41-50 | M | 18 | 3271.0 | 20586.5 | 6.3 | critical |
| P05*1 | 61-70 | M | 29 | 1029.0 | 12728.0 | 12.4 | critical |
| P06*1 | 81-90 | M | 16 | 3959.0 | 21184.0 | 5.4 | critical |
| P07*1 | 71-80 | M | 20 | 14990.0 | 36627.7 | 2.4 | critical |
| P08*1 | 21-30 | F | 14 | 212.6 | 1351.7 | 6.4 | mild |
| P09*1 | 71-80 | M | 10 | 2174.0 | 3449.9 | 1.6 | moderate |
| P10*1 | 31-40 | F | 13 | data missing*3 | | | moderate |
| P11 | 21-30 | F | 49 | 268.0 | 1051.5 | 3.9 | mild |
| P12 | 51-60 | F | 49 | 331.9 | 4815.4 | 14.5 | moderate |
| P13 | 21-30 | F | 49 | 176.1 | 866.7 | 4.9 | mild |
| P14 | 21-30 | F | 49 | 328.0 | 1355.8 | 4.1 | mild |
| P15 | 21-30 | F | 63 | 129.3 | 1185.2 | 9.2 | mild |
| P16 | 31-40 | F | 49 | 382.4 | 2198.7 | 5.7 | moderate |
| P17 | 51-60 | F | 49 | 374.8 | 5433.1 | 14.5 | mild |

| | | | | | | | |
|---|---|---|---|---|---|---|---|
| P18 | 21-30 | F | 49 | 208.0 | 3470.2 | 16.7 | mild |
| P19 | 31-40 | F | 49 | 154.8 | 1195.0 | 7.7 | moderate |
| P20 | 41-50 | F | 63 | 376.6 | 5069.5 | 13.5 | moderate |

*1 Subject included in the epitope analysis

*2 WHO COVID-19 Clinical management: living guidance, 25 January 2021, accessed 30 June 2021

*3 Not sufficient serum volume could be obtained for these serological markers.

anti-RBD IgG; unit, AU/mL; Abbott SARS-CoV-2 IgG II Quant assay
anti-N IgG; unit, Index; Abbott SARS-CoV-2 IgG assay
