## Supplement Table 2. for "High resolution linear epitope mapping of the receptor binding domain of SARS-CoV-2 spike protein in COVID-19 mRNA vaccine recipients"

Supplemental Table 2. Sequence of the peptides used in this study.

| Peptide No. | Sequence |
|---|---|
| 1 | RVQPTESIVRFPNIT |
| 2 | PTESIVRFPNITNLC |
| 3 | SIVRFPNITNLCPFG |
| 4 | RFPNITNLCPFGEVF |
| 5 | NITNLCPFGEVFNAT |
| 6 | NLCPFGEVFNATRFA |
| 7 | PFGEVFNATRFASVY |
| 8 | EVFNATRFASVYAWN |
| 9 | NATRFASVYAWNRKR |
| 10 | RFASVYAWNRKRISN |
| 11 | SVYAWNRKRISNCVA |
| 12 | AWNRKRISNCVADYS |
| 13 | RKRISNCVADYSVLY |
| 14 | ISNCVADYSVLYNSA |
| 15 | CVADYSVLYNSASFS |
| 16 | DYSVLYNSASFSTFK |
| 17 | VLYNSASFSTFKCYG |
| 18 | NSASFSTFKCYGVSP |
| 19 | SFSTFKCYGVSPTKL |
| 20 | TFKCYGVSPTKLNDL |
| 21 | CYGVSPTKLNDLCFT |
| 22 | VSPTKLNDLCFTNVY |
| 23 | TKLNDLCFTNVYADS |
| 24 | NDLCFTNVYADSFVI |
| 25 | CFTNVYADSFVIRGD |
| 26 | NVYADSFVIRGDEVR |
| 27 | ADSFVIRGDEVRQIA |
| 28 | FVIRGDEVRQIAPGQ |
| 29 | RGDEVRQIAPGQTGK |
| 30 | EVRQIAPGQTGKIAD |
| 31 | QIAPGQTGKIADYNY |
| 32 | PGQTGKIADYNYKLP |
| 33 | TGKIADYNYKLPDDF |
| 34 | IADYNYKLPDDFTGC |
| 35 | YNYKLPDDFTGCVIA |
| 36 | KLPDDFTGCVIAWNS |
| 37 | DDFTGCVIAWNSNNL |
| 38 | TGCVIAWNSNNLDSK |
| 39 | VIAWNSNNLDSKVGG |
| 40 | WNSNNLDSKVGGNYN |
| 41 | NNLDSKVGGNYNYLY |
| 42 | DSKVGGNYNYLYRLF |
| 43 | VGGNYNYLYRLFRKS |
| 44 | NYNYLYRLFRKSNLK |
| 45 | YLYRLFRKSNLKPFE |
| 46 | RLFRKSNLKPFERDI |
| 47 | RKSNLKPFERDISTE |
| 48 | NLKPFERDISTEIYQ |
| 49 | PFERDISTEIYQAGS |
| 50 | RDISTEIYQAGSTPC |
| 51 | STEIYQAGSTPCNGV |
| 52 | IYQAGSTPCNGVEGF |
| 53 | AGSTPCNGVEGFNCY |
| 54 | TPCNGVEGFNCYFPL |
| 55 | NGVEGFNCYFPLQSY |
| 56 | EGFNCYFPLQSYGFQ |
| 57 | NCYFPLQSYGFQPTN |
| 58 | FPLQSYGFQPTNGVG |
| 59 | QSYGFQPTNGVGYQP |
| 60 | GFQPTNGVGYQPYRV |
| 61 | PTNGVGYQPYRVVVL |
| 62 | GVGYQPYRVVVLSFE |
| 63 | YQPYRVVVLSFELLH |
| 64 | YRVVVLSFELLHAPA |
| 65 | VVLSFELLHAPATVC |
| 66 | SFELLHAPATVCGPK |
| 67 | LLHAPATVCGPKKST |
| 68 | APATVCGPKKSTNLV |
| 69 | TVCGPKKSTNLVKNK |
| 70 | GPKKSTNLVKNKCVN |
| 71 | PKKSTNLVKNKCVNF |
| Single mutation containing peptides | |
| | N417N |
| 29 | RGDEVRQIAPGQTGN |
| 30 | EVRQIAPGQTGNIAD |
| 31 | QIAPGQTGNIADYNY |
| 32 | PGQTGNIADYNYKLP |
| 33 | TGNIADYNYKLPDDF |
| | N417T |
| 29 | RGDEVRQIAPGQTGT |
| 30 | EVRQIAPGQTGTIAD |
| 31 | QIAPGQTGTIADYNY |
| 32 | PGQTGTIADYNYKLP |
| 33 | TGTIADYNYKLPDDF |
| | E484K |
| 52 | IYQAGSTPCNGVKGF |
| 53 | AGSTPCNGVKGFNCY |
| 54 | TPCNGVKGFNCYFPL |
| 55 | NGVKGFNCYFPLQSY |
| 56 | KGFNCYFPLQSYGFQ |
| | N501Y |
| 57 | NCYFPLQSYGFQPTY |
| 58 | FPLQSYGFQPTYGVG |
| 59 | QSYGFQPTYGVGYQP |
| 60 | GFQPTYGVGYQPYRV |
| 61 | PTYGVGYQPYRVVVL |
