## Supplement Table 3. for "High resolution linear epitope mapping of the receptor binding domain of SARS-CoV-2 spike protein in COVID-19 mRNA vaccine recipients"

Supplemental Table 3. Raw signal intensities (blank signals are subtracted).

| raw-bl | V02 | V05 | V06 | V07 | V08 | P01 | P02 | P03 | P04 | P05 | P06 | P07 | P08 | P09 | P10 |
|---|---|---|---|---|---|---|---|---|---|---|---|---|---|---|---|
| 1 | 5898.755 | 11736.7 | 3502.173 | 4159.933 | 16225.4 | 1051.194 | 59.02 | 2425.419 | 1.929 | 176.222 | 131.111 | -4.778 | 41.656 | 15.889 | 4.333 |
| 2 | 1350.664 | 6681.4 | 1729.264 | 1373.111 | 7120.8 | 162.031 | 12.223 | 238.667 | -5.06 | 54.445 | 96.267 | -4.222 | 584.513 | 8.889 | -2.889 |
| 3 | 4975.265 | 8967.7 | 1907.9 | 2725.666 | 12677.1 | 25.767 | 16.111 | 83.222 | 11.495 | 88.778 | 135.322 | 0.889 | 17.677 | 14.666 | 2.889 |
| 4 | 2754.3 | 8186.6 | 2428.627 | 1970.777 | 11423.3 | 32.011 | 15.667 | 209.667 | 121.374 | 603.223 | 283.856 | -3.888 | 5.283 | 324.556 | -0.222 |
| 5 | 2092.24 | 4444.8 | 1547.809 | 2309.733 | 12655 | 22.02 | 18.555 | 69.475 | 232.293 | 156.667 | 341.511 | -6.111 | 8.475 | 146.361 | -1.333 |
| 6 | 1549.664 | 4037.4 | 1515.173 | 2754.444 | 16892.9 | 15.262 | 23 | 27.989 | 121.989 | 538.111 | 1184.744 | -3.334 | 24.899 | 27.444 | 1.333 |
| 7 | 640.8456 | 760.8 | 987.7179 | 3153.969 | 11193.9 | 43.949 | 23.889 | 12.495 | 150.677 | 1520.889 | 946.422 | -0.556 | 67.586 | 26 | 4.889 |
| 8 | -241.063 | 346.4 | 1326.173 | 3226.666 | 10198.6 | 11.798 | 8.667 | 16.616 | 126.707 | 795.222 | 152.189 | 14 | 12.071 | 3.222 | 25.889 |
| 9 | 75.29957 | 2671.5 | 1294.355 | 2029.111 | 9290.7 | 230.273 | 50.333 | 43.636 | 229.1 | 803.667 | 213 | 110.556 | 19.636 | 59.333 | 84.333 |
| 10 | 783.6636 | 2775.9 | 3155.627 | 2308.111 | 9684.2 | 48.717 | 11 | 115.136 | 421.444 | 986.444 | 345.044 | 79.333 | 77.808 | 292.444 | 30.777 |
| 11 | 565.5726 | 4727.5 | 1831.082 | 1770 | 6061.6 | 70.353 | 56.777 | 223.829 | 442.262 | 1359.444 | 273.744 | 101.777 | 26.171 | 150.444 | 7.888 |
| 12 | 2086.573 | 7081.68 | 668.8999 | 1886.333 | 5961.1 | 521.278 | 50.778 | 111.596 | 1345.869 | 1190.111 | 1404.378 | 1720.778 | 15.869 | 34.111 | 7.111 |
| 13 | 4647.755 | 9963.40 | 1550.718 | 2269.444 | 7744.2 | 655.762 | 77.889 | 81.697 | 862.083 | 2012.333 | 1528.19 | 554.333 | 37.904 | 57.111 | 4.889 |
| 14 | 1086.573 | 4081.68 | -57.0091 | 1769.889 | 5955.4 | 509.142 | 54.889 | 78.596 | 1265.323 | 1897.445 | 797.378 | 291 | 20.596 | 218.889 | 8.334 |
| 15 | 2692.3 | 5300.4 | 1196.718 | 220.3333 | 5703.7 | 78 | 2.111 | 258.969 | 10.515 | 104.444 | 131.333 | -6.667 | 4.606 | 36.444 | 2.777 |
| 16 | 2582.3 | 7162.1 | 1384.446 | 1027.777 | 7176.4 | 72.678 | 126.861 | 3853.064 | 35.178 | 73.445 | 455.667 | 12.889 | 33.678 | 15.778 | 8.778 |
| 17 | 790.2726 | 4547.9 | 453.0819 | 586.6663 | 4360.5 | 18.856 | 3.778 | 496.402 | 38.778 | 39.112 | 235 | -4.111 | 10.829 | 10.334 | -0.444 |
| 18 | 440.4816 | 3810.4 | 1532.627 | 1619.333 | 5671.8 | 33.478 | 6.111 | 1892.547 | 321.945 | 195.222 | 310.978 | -0.111 | 5.233 | 39.945 | 4.222 |
| 19 | 2945.573 | 4096.4 | 1838.809 | 3424.333 | 12593.7 | 75.333 | 83.222 | 239.717 | 105.844 | 275 | 615.044 | 6.333 | 11.535 | 39.694 | 12.694 |
| 20 | 1009.3 | 4528.9 | 1680.264 | 2497.444 | 11885.3 | 64.411 | 62.667 | 35.929 | 105.986 | 310.333 | 672.411 | 0.667 | 22.111 | 17.555 | 3.555 |
| 21 | -413.245 | -156.5 | 273.2639 | 793.2223 | 6931.4 | 166.677 | 112.639 | 109.222 | 128.858 | 413.389 | 241.822 | 4.333 | 7.586 | 11.889 | 13.444 |
| 22 | -913.245 | 184.6 | 306.8999 | 921.8893 | 5217.6 | 5.811 | 5.778 | 5.747 | 112.475 | 745.528 | 101.411 | -0.556 | -1.434 | -1.778 | 7.667 |
| 23 | -590.245 | -556.1 | 115.4459 | 2383.222 | 4072.1 | 17.678 | 13.667 | 1.687 | 859.349 | 525.556 | 932.361 | 71.556 | 3.869 | 32.111 | -2.778 |
| 24 | -22.0634 | -210.1 | 890.2639 | 1027.889 | 3273.6 | 44.011 | 56.333 | 24.566 | 481.194 | 835.361 | 319.611 | 12.222 | 26.566 | 106.778 | 2.222 |
| 25 | 1029.664 | 2737.1 | 1561.809 | 1963 | 3496.4 | 241.1 | 50.778 | 319.909 | 409.273 | 1208 | 939.778 | 88.444 | 7.545 | 155.889 | 7 |
| 26 | 1917.823 | 3163.8 | 335.1729 | 1712.777 | 9629.1 | 1082.079 | 235.666 | 7295.389 | 4525.586 | 9002.068 | 1748.922 | 2271 | 16.313 | 168.222 | 16.778 |
| 27 | 3878.406 | 7128.6 | -70.4641 | 8509.976 | 16659.01 | 12103.16 | 5406.706 | 370 | 30378.3 | 25759.35 | 21583 | 30366.44 | 455.067 | 247.111 | -28.556 |
| 28 | 4155.073 | 11292.5 | 1004.536 | 3399.111 | 7624.8 | 1007.578 | 368.667 | 70.596 | 11428.14 | 5326.889 | 1499.278 | 4860.111 | 29.414 | 207.778 | 1.556 |
| 29 | 3932.755 | 9894.7 | 1594.718 | 2182.788 | 7861.5 | 66.844 | 7.333 | 595.717 | 34.899 | 89.555 | 252.544 | 7.333 | 34.044 | 12.555 | -7 |
| 30 | 3599.3 | 8319.0 | 893.6269 | 1149.555 | 7451.009 | 100.589 | 25.333 | 776.162 | 1113.356 | 118.667 | 1204.525 | 52.043 | 253.089 | 88.556 | 4.445 |
| 31 | 674.0276 | 4485.7 | 583.0819 | 160.0003 | 1701.1 | 543.361 | 81.384 | 1247.02 | 2065.911 | 300.111 | 1364.838 | 85.202 | 94.211 | 62 | 18.111 |
| 32 | 767.6636 | 3662.1 | 673.4459 | 432.0003 | 4726.2 | 88.678 | 15.445 | 248.323 | 365.778 | 482.111 | 588.778 | 16.667 | 8.878 | 60 | 14 |

| | | | | | | | | | | | | | | | |
|---|---|---|---|---|---|---|---|---|---|---|---|---|---|---|---|
| 33 | 1575.209 | 3954.1 | 1644.718 | 1247 | 7179.4 | 1100.692 | 363.214 | 283.909 | 797.462 | 574.778 | 1407.636 | 797.643 | 60.6 | 421.917 | 230.643 |
| 34 | -123.427 | 2180.4 | 565.0819 | 626.4443 | 5718.4 | 135.078 | 265.695 | 55.233 | 2872.153 | 166.222 | 466.978 | 69.334 | 16.078 | 362.086 | 13.667 |
| 35 | -348.154 | 751.2 | 96.0819 | 179.6663 | 2939.4 | 36.944 | 27.222 | 13.262 | 210.08 | 216.888 | 292.694 | -6.445 | 25.319 | 7.555 | 11.777 |
| 36 | -590.609 | 470.1 | 711.8999 | 883.5553 | 587.5 | 81.322 | 23.889 | 7.131 | 143.805 | 735.222 | 448.922 | -0.778 | 10.122 | 13.778 | 12.111 |
| 37 | 361.2086 | 640.3 | 352.6269 | 1940 | 888.6 | 9.789 | 7.667 | 14.253 | 139.778 | 157.667 | 171.589 | 12.778 | -2.011 | 68.778 | -1.667 |
| 38 | -312.7 | 683.2 | 218.1729 | 913.2223 | 613.3 | 39.689 | 6.778 | 62.525 | 95.389 | 524 | 572.289 | 9.889 | 21.989 | 194.333 | -0.111 |
| 39 | 2157.573 | 3845.4 | 2801.173 | 1727.889 | 6165.8 | 854.889 | 34.556 | 421.556 | 267.465 | 11821.13 | 5040.973 | 152.181 | 33.356 | 738.156 | 31.556 |
| 40 | -109.7 | 2167.4 | 1010.082 | 2234.777 | 5528 | 3764.19 | 125.889 | 414.583 | 1179.969 | 1635.333 | 17682.9 | 663.889 | -0.167 | 203.111 | 13.444 |
| 41 | 632.4816 | 2833.8 | -50.9181 | 1625.333 | 5622.1 | 961.653 | 296.378 | 239.778 | 764.278 | 1108.445 | 5588.687 | 1076.978 | 19.078 | 133.111 | 11 |
| 42 | 1744.482 | 4911.9 | 765.4459 | 2842.555 | 7189.9 | 187.233 | 88.333 | 28.424 | 2123.878 | 642.111 | 353.533 | 740.444 | 11.133 | 34.555 | 1.889 |
| 43 | 582.8456 | 6257.2 | 796.4459 | 574.8893 | 2949.8 | 17.433 | 6.777 | 72.433 | 40.242 | 78.555 | 87.333 | -1.556 | 11.333 | 19.444 | 5.777 |
| 44 | 1549.3 | 6526.9 | 572.8999 | 760.4443 | 2503.8 | 16.767 | 10.778 | 119.367 | 130.042 | 112.223 | 221.334 | 0.667 | 22.567 | 36.445 | 0.556 |
| 45 | 1567.209 | 5797.1 | 1088.809 | 1602.889 | 7512.4 | 36.236 | 63.778 | 289.911 | 128.222 | 526.778 | 279.411 | -3.111 | 10.911 | 59.444 | 18.333 |
| 46 | 3622.573 | 9249.8 | 883.4459 | 3854.788 | 12727.46 | 167.933 | 86.151 | 388.333 | 85.424 | 435.666 | 411.633 | 35.222 | 13.933 | 138.111 | 14.111 |
| 47 | 5220.028 | 11884.5 | 4496.173 | 5500.555 | 17363.4 | 357.542 | 44.81 | 221.867 | 333.167 | 304.445 | 1420.834 | 58.238 | 27.667 | 523.205 | 18 |
| 48 | -353.882 | 1395.7 | 640.7179 | 211.5553 | 1770.2 | 99.144 | 38.555 | 47.144 | 379.888 | 242.111 | 390.844 | 5.444 | 34.944 | 86.555 | 14 |
| 49 | 162.7546 | 4700.1 | 2929.264 | 1724.151 | 10311.1 | 711.136 | 427.944 | 171.027 | 144.444 | 722.535 | 3246.301 | 470.829 | 1496.044 | 21.555 | 873.194 |
| 50 | -516.609 | 1492.7 | 618.7179 | -339.667 | 0 | 64.022 | 421.355 | 15.622 | 70.722 | 137.778 | 222.122 | 31.555 | 50.222 | 8.889 | 66.555 |
| 51 | -553.609 | 754.5 | 1319.627 | 625.7773 | 1996 | 9.944 | 42 | 11.544 | 68.171 | 189.777 | 113.544 | 21.555 | 8.044 | 19 | 4.333 |
| 52 | 80.14357 | 2135.4 | 1138.809 | 783.4443 | 5376.1 | 30.022 | 25.555 | 26.422 | 36.313 | 660.444 | 160.622 | 102.778 | 4.522 | 55.333 | -1.778 |
| 53 | 377.6636 | 2037.9 | -38.8271 | 593.7773 | 4025.1 | 324.653 | 7.667 | 21.378 | 84.869 | 485.889 | 864.334 | 1952.578 | -2.522 | 29.207 | 3.889 |
| 54 | 1030.209 | 5487.6 | 193.5359 | 1436.889 | 7255.4 | 188.556 | 24.667 | 45.578 | 224.323 | 346.111 | 1860.111 | 168.334 | 5.078 | 24.778 | 3.889 |
| 55 | 1060.028 | 5634.9 | 1004.991 | 849.4443 | 5694.8 | 238.333 | 35.556 | 30.4 | 278.833 | 316.667 | 905.1 | 152.889 | 9.3 | 17.222 | 14.222 |
| 56 | 1939.482 | 6718.9 | 936.4459 | 1728.666 | 6976.6 | 86.133 | 14.889 | 13.833 | 303.333 | 366.555 | 183.333 | 82.777 | 6.633 | 20.444 | 2.111 |
| 57 | 1858.755 | 7426.6 | 1230.446 | 1594.444 | 3266.7 | 15.178 | 7.778 | 21.978 | 4.051 | 216.445 | 103.678 | -8.111 | 2.278 | -0.444 | 3.667 |
| 58 | 3233.028 | 7517.6 | 1334.718 | 3192.222 | 4397.555 | 17.522 | 21.444 | 122.522 | 17.222 | 1050.949 | 1164.858 | 1.222 | 6.022 | 28.889 | 8.778 |
| 59 | 1983.755 | 8075.6 | 852.2639 | 1949.333 | 5918.2 | -11.333 | -2.666 | 39.167 | 62.212 | 1184.485 | 242.889 | 55.667 | -27.633 | -8.889 | -26.889 |
| 60 | 1525.209 | 7869.0 | 1421.536 | 2826 | 6767.5 | 43.944 | 16.111 | 45.666 | 91.353 | 213.222 | 232.144 | 4.555 | 2.744 | 26.444 | 7.444 |
| 61 | 1095.3 | 8340.6 | 648.5359 | 2039.444 | 3357.3 | 107.856 | 37.667 | 54.156 | 74.101 | 3454.623 | 228.985 | -0.222 | 15.556 | 31.223 | 6.889 |
| 62 | 445.4816 | 5717.5 | 814.7179 | 1379.333 | 3560.2 | 73.822 | 130.555 | 41.922 | 150.767 | 4060.495 | 534.222 | 0.666 | 23.722 | 35.555 | 87.778 |
| 63 | 1116.209 | 3735.2 | 1487.627 | 555.7773 | 1898.6 | 50.222 | 21.333 | 51.347 | 114.022 | 279.222 | 475.936 | 24.597 | 34.793 | 17.222 | 47.055 |
| 64 | 1059.3 | 5344.5 | 1845.809 | 516.4443 | 3412.8 | 27.878 | 14.778 | 25.178 | 109.653 | 130.556 | 180.378 | 10.778 | 12.178 | 11.111 | 18.667 |
| 65 | 261.1176 | 6324.9 | 2961.082 | 846.0003 | 7649.3 | 7.433 | 12.111 | 8.633 | 60.904 | 196.333 | 132.133 | -1.667 | 0.533 | 9.555 | 3.889 |
| 66 | 4106.973 | 5840.6 | 7496.991 | 3843.222 | 17577.37 | 39.233 | 52.666 | 158.633 | 73.878 | 361.555 | 139.133 | 10.555 | -5.467 | 2.222 | -5.556 |

| | | | | | | | | | | | | | | | |
|---|---|---|---|---|---|---|---|---|---|---|---|---|---|---|---|
| 67 | 3310.573 | 14211.8 | 4998.355 | 5835.111 | 15127.4 | 44.056 | 42.223 | 65.256 | 83.011 | 163.667 | 222.856 | 67 | -1.344 | 34.889 | 1.112 |
| 68 | 2791.664 | 14216.4 | 1839.991 | 6243.06 | 17157.37 | 86.433 | 52.666 | 39.133 | 90.788 | 196.111 | 534.333 | 38.777 | 3.933 | 16.222 | 2.222 |
| 69 | 4669.573 | 17842.9 | 2764.082 | 4012 | 11441.7 | 297.667 | 94.223 | 93.267 | 115.849 | 592 | 566.267 | 33.778 | 8.667 | 29.223 | 6.556 |
| 70 | 3448.482 | 17271.2 | 3651.991 | 6563.633 | 8992.2 | 97.078 | 50.153 | 48.578 | 163.323 | 727.778 | 231.178 | 19.556 | 13.178 | 104.921 | 3.778 |
| 71 | 4970.937 | 10457.1 | 2181.082 | 4219.666 | 5655.6 | 18.364 | 16.222 | 36.4 | 21.364 | 134.889 | 95.727 | -2.556 | 3.2 | 8.111 | -3.111 |
| N417N | | | | | | | | | | | | | | | |
| 29 | 4217.118 | 10766.68 | 2788.627 | 2386.111 | 8558.017 | 48.142 | 40 | 52.978 | 130.051 | 194.556 | 871.778 | 9.222 | 7.478 | 52.667 | 0.222 |
| 30 | 3562.664 | 13991.4 | 2955.173 | 6844.666 | 11278.93 | 28.525 | 54.222 | 156.289 | 188.162 | 341.445 | 341.889 | 291.756 | -2.611 | 50.111 | 6.111 |
| 31 | 761.7546 | 8268.675 | 1232.173 | 3159.444 | 3155.1 | 64.707 | 290 | 54.589 | 380.351 | 676.222 | 378.98 | 28.333 | 11.489 | 47.222 | 41.667 |
| 32 | 1325.482 | 6500.22 | 3253.536 | 3263.889 | 7501.4 | 651.644 | 465.361 | 344.277 | 542.08 | 582 | 511.353 | 108.544 | 399.829 | 72.111 | 65.555 |
| 33 | 1867.118 | 10858.58 | 3653.809 | 3548.444 | 6614.3 | 268.264 | 407.434 | 117.889 | 690.822 | 340.445 | 331.344 | 5.778 | 23.789 | 120.735 | 34.556 |
| N417T | | | | | | | | | | | | | | | |
| 29 | 2757.573 | 7307.402 | 3687.718 | 3254.111 | 11435.4 | 62.485 | 109.223 | 56.167 | 55.303 | 517.556 | 478.122 | 8.556 | 7.867 | 21.334 | 12.445 |
| 30 | 1084.573 | 17339.77 | 9771.173 | 4062.222 | 14326.6 | 25.818 | 75.222 | 35.9 | 168.545 | 626.778 | 225.727 | 24.111 | 6.6 | 166.429 | 20.667 |
| 31 | 858.3906 | 7182.493 | 2483.355 | 3178.889 | 10349.7 | 55.838 | 404.611 | 22.011 | 123.929 | 1206 | 291.747 | 24 | 17.611 | 82.656 | 9.444 |
| 32 | 2472.664 | 7329.675 | 1902.627 | 2668.444 | 11144.1 | 373.889 | 1923.197 | 180.289 | 234.162 | 1880.778 | 964.616 | 35.445 | 147.972 | 82.556 | 5.889 |
| 33 | 2182.118 | 15713.04 | 4810.991 | 6492.444 | 8218.4 | 432.587 | 662.598 | 51.044 | 487.301 | 2005.666 | 459.808 | 6.555 | 42.744 | 516.675 | 0.222 |
| E484K | | | | | | | | | | | | | | | |
| 52 | 2924.846 | 9065.675 | 2575.355 | 3591.333 | 16943.6 | 83.08 | 50.888 | 32.544 | 40.626 | 383.444 | 105.044 | -8.667 | -2.056 | -8.778 | 2.333 |
| 53 | 2507.209 | 7142.402 | 2184.9 | 3950.444 | 12114.1 | 38.889 | 77.889 | 1061.489 | 19.707 | 221.445 | 132.489 | 12.389 | -0.011 | 12.667 | 15.222 |
| 54 | 3428.118 | 7688.13 | 4884.991 | 3585.444 | 9925.6 | 55.273 | 252.556 | 68.9 | 53 | 347.111 | 205.5 | 2 | 11.1 | 6.222 | 9.111 |
| 55 | 3128.937 | 8151.22 | 4931.173 | 4643.889 | 12089.6 | 97.079 | 206.666 | 43.222 | 107.131 | 421.222 | 104.522 | -1.222 | 43.079 | 11.555 | -0.111 |
| 56 | 2383.391 | 8093.039 | 4252.355 | 4041.111 | 11561.4 | 148.445 | 139.889 | 33.278 | 64.403 | 760.111 | 152.878 | -2.889 | 15.178 | 16.889 | 2.889 |
| N501Y | | | | | | | | | | | | | | | |
| 57 | 2262.209 | 7069.948 | 5450.264 | 3454.555 | 13795.2 | 135.292 | 159.467 | 11.967 | 39.167 | 576.778 | 224.567 | -8.111 | -1.333 | 3 | -1.666 |
| 58 | 2989.573 | 7605.311 | 4383.627 | 3898.222 | 9998.8 | 122.051 | 247.978 | 45.445 | 56.678 | 525.111 | 626.028 | 0.222 | 9.278 | 10.222 | 11.222 |
| 59 | 3274.482 | 5450.402 | 3166.446 | 3356.333 | 13039.2 | 99.778 | 222.078 | 25.978 | 101.505 | 688.334 | 390.378 | 8.889 | 2.478 | 42.889 | 37.445 |
| 60 | 2696.209 | 7658.039 | 3545.264 | 4425.444 | 17699.2 | 59.929 | 136.333 | 31.311 | 64.747 | 935.444 | 200.411 | -3.667 | 1.911 | 95.111 | 4.667 |
| 61 | 1722.846 | 7234.948 | 4839.718 | 4558.111 | 10792.3 | 116.04 | 200.555 | 28.422 | 117.04 | 4042.389 | 187.122 | 0.111 | 8.522 | 38.111 | -2.334 |
